# Genetic Contribution of Cardiorespiratory Fitness in Morbidity and Mortality: A Prospective FinnGen and HUNT study

**DOI:** 10.1101/2025.06.25.25330268

**Authors:** L Joensuu, V Lukander, P Herranen, NP Tynkkynen, U Kujala, R Lopéz-Bueno, AN Nordeidet, M Klevjer, K Øvretveit, U Wisløff, A Bye, U Ekelund, M Ollikainen, FinnGen, E Sillanpää

## Abstract

**Objectives:** To quantify the contribution of cardiorespiratory fitness (CRF) genetics in common non-communicable disease (NCD) and mortality risk and to assess whether health discrepancies exist between “inherited” and “gained” CRF.

**Methods:** We used a validated SBayesR-based genome-wide polygenic score, leveraging information from 905,707 single-nucleotide polymorphisms, to measure CRF genetics (PGS CRF). Associations with register-based incident cardiovascular disease, cancer, pulmonary disease, type 2 diabetes (T2D), and all-cause mortality were analysed using Cox proportional hazards models in the FinnGen cohort (N=262,137; 53.5-y at baseline, 52.0% women) and replicated in the HUNT3 cohort (N=26,115; 59.0-y, 52.4% women). In HUNT3, we also compared the health characteristics and disease risk of individuals having age- and sex-specific high CRF (V̇O_2max_) and high PGS CRF (group “inherited” CRF) to those having high CRF but low PGS CRF (group “gained” CRF), N=375 vs. 279, respectively.

**Results:** Higher PGS CRF was associated with lower risk of lung cancer (hazard ratio [HR] 0.95, 95% confidence interval 0.93–0.98), chronic obstructive pulmonary disease (HR 0.98, 0.96–1.00), T2D (HR 0.98, 0.96–0.99), and all-cause mortality (HR 0.98, 0.98–0.99) in the most adjusted model per each standard deviation increase in PGS. In sensitivity analyses including never-smokers, the association between PGS CRF and T2D remained statistically significant. Replication analyses supported main observations. No differences in health outcomes were observed between individuals with “inherited” and “gained” CRF.

**Conclusions:** PGS CRF requires further development, but the findings suggest genetic predisposition accounts for some, albeit a limited proportion, of the public health benefits observed with CRF.

**Summary box:** *What is already known on this topic:* Cardiorespiratory fitness (CRF) is a well-known correlate of health and longevity, with an expected strong genetic component. However, it is not known to what extent genetics explain the health benefits of CRF or if “inherited” CRF is more health-protective than CRF “gained” through exercise.

*What this study adds:* We found that current polygenic metrics for CRF show modest protective associations against type 2 diabetes, but not against cardiovascular disease, cancers, pulmonary disease or all-cause mortality after controlling for potential covariates. We did not observe differential associations between “inherited” and “gained” CRF with health outcomes.

*How this study might affect research, practice or policy:* Genetic confounding is expected to play a limited role in the relationship between CRF and the risk of common NCDs and mortality. Therefore, promotion of CRF remains a viable public health strategy.

## Introduction

Physical fitness is a set of attributes that people have or achieve and which relate to their ability to perform physical activity.^1^ Cardiorespiratory fitness (CRF) is the ‘integrated ability to transport oxygen from the atmosphere to the mitochondria to perform physical work’^2^ and is among the most studied aspects of physical fitness. CRF is associated with non-communicable diseases (NCDs) throughout the lifespan.^2–4^ The world’s leading NCDs include heart disease, cancer, chronic respiratory disease, and diabetes.^5^ A recent umbrella review with 20.9 million individual observations showed that better CRF is systematically associated with a lower risk of incident hypertension, heart failure, stroke, atrial fibrillation, type 2 diabetes (T2D), and cardiovascular, sudden cardiac, all-cancer, and lung cancer mortality.^6^ In quantitative estimates, one metabolic equivalent of task (1-MET) increase in CRF was associated with a lower risk of cardiometabolic diseases (disease-specific hazard ratios [HRs] ranging from 0.82 to 0.97) and with lower risk of all-cause mortality (HRs from 0.83 to 0.89).^6^

CRF is, however, known to be strongly influenced by genetics.^7^ Meta-analysed heritability estimates derived from twin and family studies range from 44% to 68%, depending on the study and measure of CRF.^8^ Genetic correlation studies indicate shared genetic material between CRF and several NCD risk factors.^9^ Furthermore, some of the genetic variants related to CRF are expression quantitative trait loci that may influence gene expression in the heart, artery, lung, and adipose tissue.^9,10^ Hence, a favourable genotype in the context of CRF may act as the basis for better structure and function of the lungs, heart, vasculature, and cellular metabolism and influence systemic inflammation and immune function – all aspects highly involved in both CRF and common NCDs.^4,9^ However, it is not yet known to what extent genetics explain the beneficial health effects of high CRF.

It also remains unclear whether the health benefits of genetically influenced CRF versus CRF gained through adaptations to exercise differ. Studies have found mixed evidence showing that twins discordant in their CRF did not differ in baseline characteristics or in the incidence of later cardiovascular disease (CVD).^11^ However, former elite endurance athletes lived 2.4 years longer than their brothers.^12^ Clarifying whether the health benefits of CRF differ based on whether it is genetically influenced or acquired through exercise may provide insight into the extent of genetic confounding in previous research.

Genome-wide polygenic scores (PGSs) can be used to estimate an individual’s genetic makeup more accurately than was previously possible.^13^ In this study, we developed and applied a PGS to quantify the association between CRF genetics and the risk of major NCDs and mortality. We furthermore evaluated differences in health characteristics and disease risk among individuals with similarly high measured CRF but differing PGSs. We assumed that in the presence of high CRF but low genetic susceptibility to high CRF, CRF is “gained” over time. Conversely, when high measured CRF coincides with genetic susceptibility to high CRF, CRF is “inherited.” We hypothesised that 1) higher PGS CRF is associated with lower risk of incident CVD, cancer, chronic respiratory disease, T2D, and all-cause mortality and conservatively that 2) “inherited” and “gained” CRF show similar magnitudes of association.

## Methods

The study design is illustrated in Figure 1. We utilised the prospective FinnGen cohort for the main analysis and HUNT3 for validation, replication, and group comparisons (Figure 1). We also conducted additional sensitivity analyses in FinnGen and exploratory analyses in HUNT3. This study is reported in accordance with the STROBE^14^ and CHAMP statements.^15^

**Figure 1.**
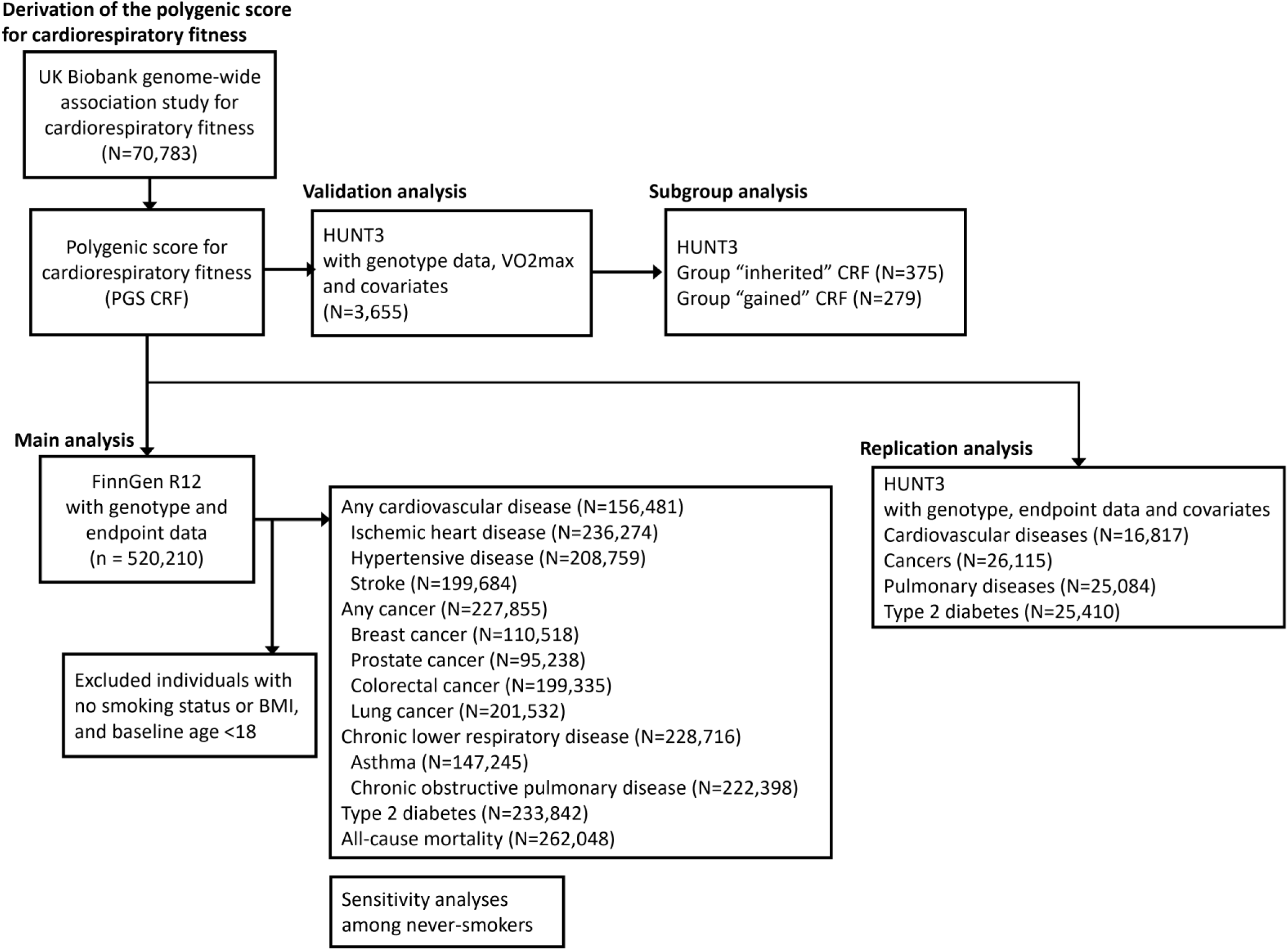
Study design and flowchart.

### Study cohorts

In short, FinnGen is an ongoing Finnish public–private partnership research project linking genotype and digital health registry data using personal identification numbers.^16^ The recruitment is operated mainly via healthcare contacts. FinnGen data release 12 comprises 520,210 participants (∼9.4% of the Finnish population) (https://www.finngen.fi/en).

HUNT (the Trøndelag Health Study) is a large, population-based cohort study conducted in the Trøndelag region of Norway integrating health survey data, biological samples, and national health registries.^17,18^ The study includes one of the largest available datasets for CRF measured by indirect calorimetry. Since its inception in 1984, phenotypic and genetic data have been collected in several waves (HUNT1–HUNT4, https://www.ntnu.edu/hunt).^19^ A total of 50,800 individuals aged 20 years and older participated to HUNT3 in 2006–2008.

### Outcomes

The selection of NCDs was based on the World Health Organization’s criteria for the most common NCDs.^5^ We also used all-cause mortality as an outcome. FinnGen data originate from the Care Register for Health Care (available from 1968), Finnish Cancer Registry (1953), Causes of Death (1969), and Finnish Social Insurance Institution Drug Purchase registers (1995). Any CVD, with subcategories of ischaemic heart diseases, hypertension, and stroke; any cancer (subcategories: breast, prostate, colorectal, and lung); chronic lower respiratory disease (subcategories: asthma and chronic obstructive pulmonary disease [COPD]), and T2D were used as outcomes. The selection of the subcategories was based on the most prevalent endpoints in the FinnGen R12 cohort. Relevant ICD-10 codes and FinnGen labels for data browsing are available in Table S1. We utilised the same ICD codes in the HUNT replication analyses. The HUNT data are from the Nord-Trøndelag Health Trust discharge register (available from 1987).

### Exposure

We chose the largest genome-wide association study (GWAS) for CRF available at the time of the analysis as the base data for PGS CRF after careful evaluation.^9^ This publicly available summary statistic indicates each single_-_nucleotide polymorphism’s (SNP’s) effect size on CRF. The effect sizes are based on information from 70,783 genotyped UK Biobank participants who had undergone a submaximal cycle ergometer test. In the GWAS, estimated maximal workload relative to body weight in kilograms (watts/kg) was selected to represent CRF. Workload and heart rate data were collected during a 6-min exercise period where participants screened to have ‘minimal’ or ‘small’ risk categories had their workload increased, and a ‘medium’ risk group kept their workload constant during the test. Maximal workload was thereafter extrapolated using age-estimated maximum heart rate. The risk category was considered a covariate in later genetic association analyses.^9^ The full test protocol is published elsewhere.^20^ The GWAS identified 12 statistically significant SNPs and multiple functionally relevant genes.^9^ The SNP heritability was 10.5% based on Complex-Traits Genetics Virtual Lab,^21^ indicating its suitability as base data for constructing a PGS.^13^

We used the SBayesR method to construct the PGS CRF for each participant.^22^ First, we retrieved the reported weights for 8,918,920 SNPs. Then, we restricted this genetic information to 1,074,896 HapMap3 SNPs to ensure computational efficiency while maintaining the genome-wide representation, focusing on common variants (minor allele frequency, MAF >5%) and good imputation coverage.^23^ To correct for weight inflation caused by linkage disequilibrium (LD), we subsequently applied an LD-reference panel consisting of 50,000 randomly selected UK Biobank participants. These procedures resulted in 916,113 SNPs and their relevant weights, which were used to calculate a PGS CRF for each participant in FinnGen and HUNT3. The full pipeline is described elsewhere.^24^ The final number of processed variants was 905,707 in FinnGen and 915,971 in HUNT3. We used a standardised PGS score in the analyses (mean of 0, standard deviation [SD] of 1).

The constructed PGS CRF showed small but statistically significant associations against the gold standard measure of CRF, measured by indirect calorimetry during a maximal graded treadmill test (Cortex MetaMax II, Cortex Biophysik GmbH, Leipzig, Germany). The PGS CRF explained 0.20% of variation in V̇O_2max_ mL/kg/min and 0.21% in V̇O_2max_ mL/kg^0.75^/min. The observed associations were comparable in magnitude to those of previously validated exercise-related polygenic scores associated with incident NCDs.^25–28^ We also tested two alternative PGSs in which 1) the SNP weights were based on a GWAS for V̇O_2max_ scaled to fat-free mass,^29^ and scoring as previously described (SBayesR method) (PGS CRF FFM), and 2) based on favorable SNPs^29^ and their summarised effect sizes (GRS CRF). These alternative scores did not, however, improve the variation explained in measured CRF and were therefore excluded from further analyses (see Table S2 for validation cohort descriptives, Table S3 for detailed results, Figure S1 for density plots, and Figure S2 for a correlation heatmap between genetic scores and selected markers of CRF).

### Genotyping, quality control, and imputation

The FinnGen samples were genotyped with Illumina and Affymetrix chip arrays (Illumina Inc., San Diego, and Thermo Fisher Scientific, Santa Clara, CA, USA). Detailed genotyping, quality control, and imputation information are available elsewhere: https://finngen.gitbook.io/documentation/. In HUNT3, standard and customized HumanCoreExome arrays from Illumina were used. Full details of genotyping, imputation, and quality control can be found in other sources.^19^

### Covariates

Covariates were selected based on prior literature to account for potential biases arising from genotyping procedures and individual characteristics. The first 10 principal components of ancestry adjust for genetic stratification.^30^ Genotyping batch was included to correct for potential systematic biases introduced by the genotyping process. Body mass index (BMI), smoking status (never, former, current), and sex were obtained from the Finnish Biobanks and HUNT and included both self-reported and measured data.^16,19^

### Patient and public involvement

The participants were not involved in forming the research questions, outcome measures, design, or other aspects of the study.

### Equity, diversity, and inclusion statement

The group of authors is gender balanced, with seven women and seven men, and includes researchers in different organisations, career stages, and disciplines. Our study populations include a balanced distribution of male and female subjects, although participants with an established connection to health services may be overrepresented in FinnGen.

### Statistical analysis

#### Survival analyses

We used Cox proportional hazard models and R package survival^31^ to estimate the HRs and 95% confidence intervals (CI) between PGS CRF and incident NCDs and mortality in the FinnGen cohort. Visual inspection of Schoenfeld residuals and log-minus-log plots indicated that the fully adjusted models satisfy proportional hazard assumptions.^32^ PGS×SEX interaction was tested, but no replicable sex differences were observed (all p_interaction_ >0.370); thus, analyses were conducted with men and women combined. We used age as the timescale. Start of follow-up was set at each participant’s baseline age (age at blood sampling) to ensure that the study mimicked the prospective designs of previous CRF literature. Follow-up ended with whichever came first among the first record of the endpoint of interest, death, or the end of follow-up on 31 December 2021. Underaged individuals (<18-y) were excluded to maintain consistency between the FinnGen and HUNT cohorts. Model 1 included sex, the first 10 principal components of ancestry, and genotyping batch. Model 2 additionally included BMI, and Model 3 additionally included smoking status. Utilising the weights of the European standard population,^33^ age-standardised incidence was calculated when relevant.

#### Sensitivity analyses

As a sensitivity analysis, we restricted the Cox proportional hazards model to never-smokers in the FinnGen study. The model specifications were otherwise identical to those used in the main survival analyses.

#### Replication analyses

The replication analyses in HUNT3 were conducted similarly to the main analyses in FinnGen, except that in the absence of mortality data, the follow-up was defined as the first record of the endpoint of interest or last verified contact with healthcare until the end of year 2024. To ensure sufficient sample size, outcomes were restricted to main disease categories: CVDs, cancer, pulmonary diseases, and T2D.

#### Group comparisons

A priori power calculations were used to determine sufficient sample size and group cut-offs (expected small effect size, power at 0.80, N>400). Groups were formed based on individuals in the ≥70^th^ percentile in age- and sex-specific V̇O_2max_. Age was handled in 5-year bins separately for both sexes. Individuals with high V̇O_2max_ and high PGS CRF (both ≥70^th^ percentile) were labelled as “inherited” CRF (Table S4). Individuals with high V̇O_2max_ but low PGS CRF (≤30^th^ percentile) were labelled as “gained” CRF. The differences between the groups in health-related characteristics were evaluated for continuous variables with ANOVA. Assumptions of normality were inspected visually and/or with Shapiro-Wilk’s test, homogeneity with Levene’s Test, and statistical differences with Tukey’s HSD test. For categorical variables, the differences were tested with the chi-squared test. Variables which considerably deviated from normal distribution (such as MET h/week and triglycerides) were square-root and log-transformed respectively. As the observed statistical associations did not differ between original and transformed values, only original values are shown. The Cox proportional hazards model was used in exploratory analyses to investigate disease incidence between the groups. In all analyses in the study, the statistical significance level was set to <0.05.

## Results

Descriptive data on all 262,137 participants in the FinnGen sample are shown in Table 1. The incidence and follow-up times were disease-specific. For example, 17.4% participants were diagnosed with any CVD (45,504 cases) with an age-standardised incidence of 354.86 per 10,000 person-years (Figure 2). The average follow-up for any CVD was 9.23 years and a total of 419,935 person-years.

**Figure 2.**
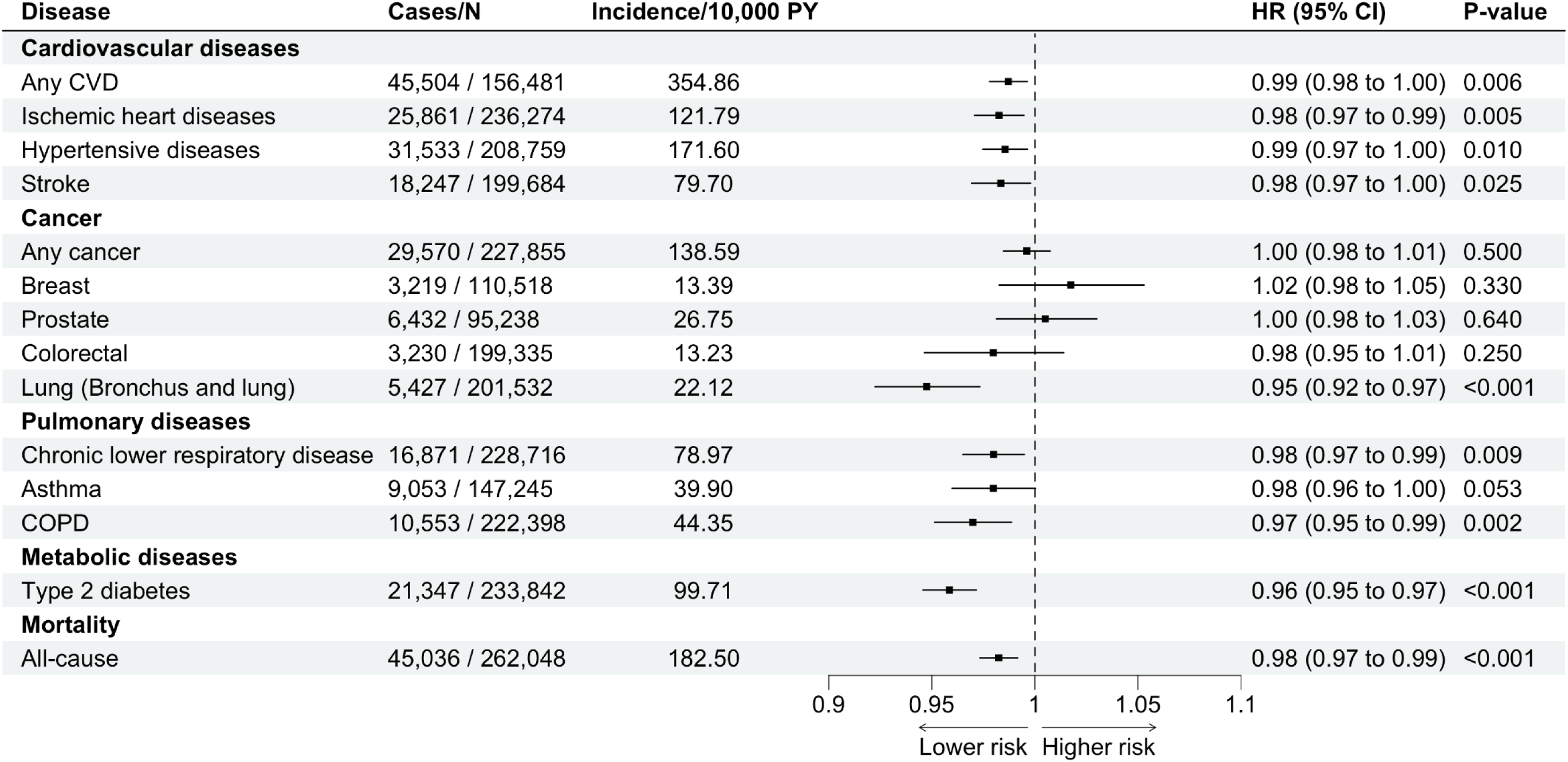
Associations of polygenic score for cardiorespiratory fitness with common non- communicable diseases and all-cause mortality before adjustment for BMI and smoking. The model was adjusted for sex, the first 10 principal components of ancestry, and genotyping batch.

**Table 1.** Baseline characteristics of participants in the FinnGen study.

| Characteristic | All<br>(n=262,137) | Women<br>(n=136,331) | Men<br>(n=125,806) |
| --- | --- | --- | --- |
| Age (y) | 53.5 (16.0) | 51.4 (16.6) | 55.8 (15.0) |
| BMI (kg/m<sup>2</sup>) | 27.4 (5.3) | 27.4 (5.8) | 27.5 (4.6) |
| Height (cm) | 170.4 (9.1) | 164.4 (6.4) | 176.9 (6.9) |
| Weight (kg) | 79.8 (17.3) | 74.01 (16.4) | 86.0 (16.1) |
| Smoking status n (%) |  |  |  |
| Never | 130,211 (49.7) | 82,797 (60.7) | 47,414 (37.7) |
| Former | 61,788 (23.6) | 28,499 (20.9) | 33,289 (26.5) |
| Current | 70,138 (26.8) | 25,035 (18.4) | 45,103 (35.9) |

Values are means (standard deviations) for continuous variables and n (%) for categorical/binary. BMI, body mass index.

### Associations between PGS CRF, morbidity, and mortality

Each SD increase in PGS CRF was associated with a reduced risk of any CVD (HR 0.99, 95% CI 0.98–1.00), ischaemic heart diseases (HR 0.98, 0.97–0.99), hypertensive diseases (HR 0.99, 0.97–1.00), and stroke (HR 0.98, 0.97–1.00) (Figure 2). Higher PGS CRF was associated with a lower risk of lung cancer (HR 0.95, 0.92–0.97) but not with other cancers. Higher PGS CRF was associated with a lower risk of chronic lower respiratory disease (HR 0.98, 0.97–0.99) and COPD (HR 0.97, 0.95–0.99) but not with asthma (HR 0.98, 0.96–1.00, p=0.053). Higher PGS CRF was associated with a lower risk of T2D (HR 0.96, 0.95–0.97) and all-cause mortality (HR 0.98, 0.97–0.99) (Figure 2).

When adjusting for BMI, relevant changes were observed in CVDs. The associations were attenuated and persisted after adjustment for BMI only in ischaemic heart diseases (HR 0.99, 0.97–1.00). For other NCDs, the statistically significant associations between PGS CRF and disease endpoints remained stable after adjusting for BMI (lung cancer, chronic lower respiratory disease, COPD, T2D, and all-cause mortality; Figure S3 in the Supplement). After additional adjustment for smoking, statistically significant associations were observed only with lung cancer (HR 0.95, 0.93–0.98), COPD (HR 0.98, 0.96–1.00), T2D (HR 0.98, 0.96–0.99), and all-cause mortality risk (HR 0.98, 0.98–0.99; Figure 3). For these statistically significant associations in the fully adjusted model, we inspected cumulative incidence among high PGS CRF (>90^th^ percentile) and low PGS CRF (<10^th^ percentile) individuals. The data indicated protective associations for high PGS CRF against all-cause mortality throughout the lifespan, while the benefits for COPD, lung cancer, and T2D emerged after midlife or in old age (Figure 4).

**Figure 3.**
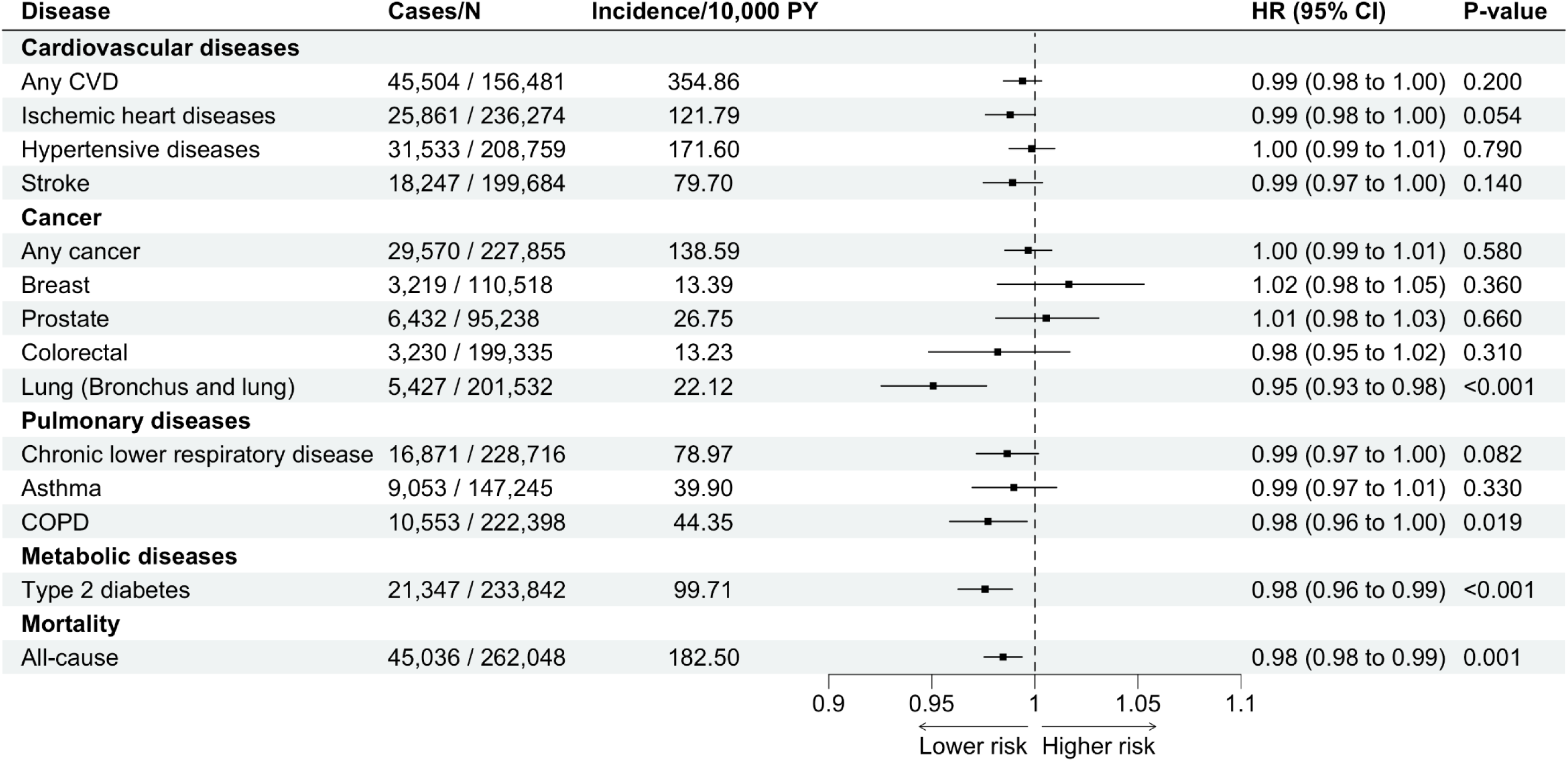
Associations of polygenic score for cardiorespiratory fitness with common non- communicable diseases and all-cause mortality in the fully adjusted model. The model was adjusted for sex, the first 10 principal components of ancestry, genotyping batch, BMI, and smoking status.

**Figure 4.**
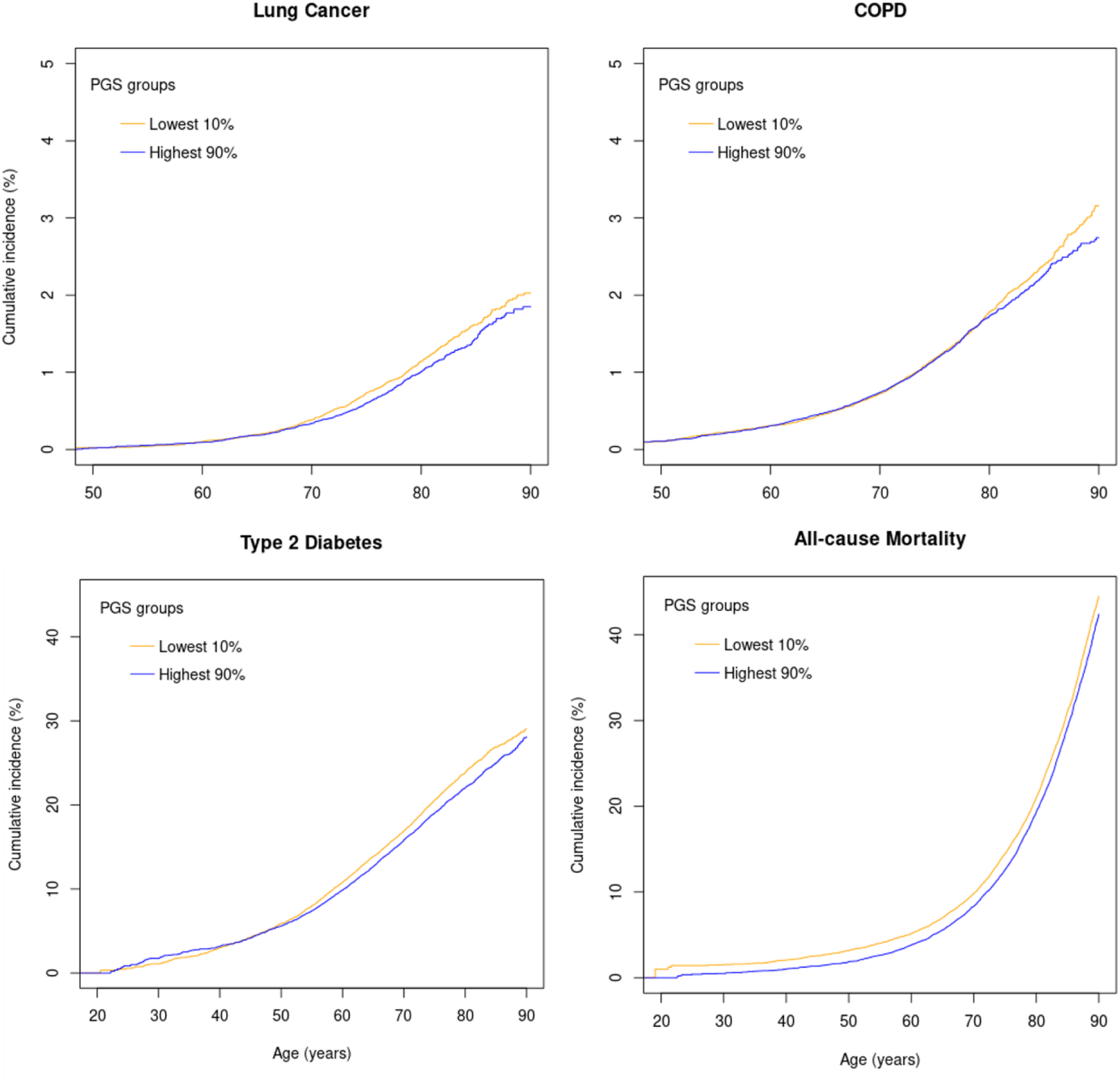
Cumulative incidence of chronic obstructive pulmonary disease, lung cancer, type 2 diabetes, and all-cause mortality in individuals with high (<90^th^ percentile) and low (<10^th^ percentile) polygenic score for cardiorespiratory fitness. Cumulative incidences are from Cox proportional hazards model and adjusted for sex, the first 10 principal components of ancestry, genotyping batch, BMI, and smoking status.

In sensitivity analyses among never-smokers, only the association with T2D remained statistically significant (Figure S4). In replication analyses (see Table S5 for sample characteristics), the association for T2D replicated in HUNT3 although attenuated after adjustment for BMI (HR 0.95, 0.92–0.99 in Model 1; HR 0.99, 0.95–1.03 in Model 2, Table S6). The associations between PGS CRF and any CVD, any cancer, and pulmonary diseases were equally as non-significant in HUNT3 as in FinnGen (HR 1.01, 0.99–1.03; HR 1.02, 0.98–1.06; and HR 0.98. 0.95–1.01, respectively, in the fully adjusted models, Table S6).

### Health discrepancies between “inherited” and “gained” CRF

The groups labelled as “inherited” CRF and “gained” CRF did not differ in health characteristics, including self-reported health, BMI, blood pressure, and blood lipids (Table 2). Overall, both groups’ characteristics were at a healthy level. Exercise habits prior to the study were unknown, but the groups did not differ in their self-reported physical activity at the time of the evaluation (Table 2). In further exploratory analyses of a subsample of individuals with health register data (N=187–232), the “gained” CRF group showed systematically lower risk estimates (HR <1.00) compared to the “inherited” CRF group for incident CVD (HR 0.83, 0.60–1.19), cancer (0.84, 0.41–1.69), pulmonary disease (HR 0.67, 0.20–2.24), and T2D (0.52, 0.05–5.90) (Table S7). However, the analyses were limited by statistical power and should be interpreted with caution.

**Table 2.** Characteristics of individuals with similar cardiorespiratory fitness but different PGS CRF.

| Characteristic | N | “Inherited” CRF |  | “Gained” CRF |  | P for difference |
| --- | --- | --- | --- | --- | --- | --- |
| | | High $\dot{V}O_{2max}$<br>High PGS CRF | N | High $\dot{V}O_{2max}$,<br>Low PGS CRF | | |
| PGS CRF (Z-score) | 375 | 1.16 (0.5) | 279 | -1.16 (0.5) |  | <0.001 |
| $\dot{V}O_{2max}$ (mL/kg/min) | 375 | 48.5 (8.6) | 279 | 48.9 (8.6) | | 0.471 |
| $\dot{V}O_{2max}$ (mL/kg<sup>0.75</sup>/min) | 375 | 141.0 (27.1) | 279 | 142.0 (27.3) | | 0.459 |
| Age (years) | 375 | 48.7 (12.4) | 279 | 47.7 (13.3) |  | 0.306 |
| Women n (%) | 375 | 208 (55.5%) | 279 | 145 (52.0%) |  | 0.419 |
| SES n (%) | 375 |  | 279 |  |  | 0.613 |
| High | 191 | 50.9% | 153 | 54.8% |  |  |
| Medium | 168 | 44.8% | 115 | 41.2% |  |  |
| Low | 16 | 4.3% | 11 | 3.9% |  |  |
| BMI (kg/m<sup>2</sup>) | 375 | 24.2 (2.7) | 279 | 24.0 (2.45) |  | 0.446 |
| Smoking status n (%) | 375 |  | 279 |  |  | 0.259 |
| Never | 222 | 59.2% | 163 | 58.3% |  |  |
| Former | 113 | 30.1% | 75 | 26.9% |  |  |
| Current | 40 | 10.7% | 41 | 14.7% |  |  |
| MET (h/week) | 375 | 15.9 (8.6) | 279 | 16.5 (9.3) |  | 0.377 |
| Self-reported health n (%) | 375 |  | 279 |  |  | 0.336 |
| Poor | 0 | 0.0% | 1 | 0.4% |  |  |
| Not so good | 16 | 4.3% | 15 | 5.4% |  |  |
| Good | 208 | 55.5% | 139 | 49.8% |  |  |
| Very good | 151 | 40.3% | 124 | 44.4% |  |  |
| Systolic blood pressure (mm/Hg) | 364 | 126 (14.9) | 267 | 126 (14.5) |  | 0.992 |
| Diastolic blood pressure (mm/Hg) | 364 | 72.0 (10.7) | 267 | 72.0 (10.2) |  | 0.986 |
| Total cholesterol (mmol/L) | 364 | 5.4 (1.01) | 267 | 5.3 (1.00) |  | 0.267 |
| HDL cholesterol (mmol/L) | 364 | 1.5 (0.36) | 267 | 1.5 (0.35) |  | 0.775 |
| LDL cholesterol (mmol/L) | 364 | 3.6 (0.9) | 267 | 3.5 (0.9) |  | 0.325 |
| Triglycerides (mmol/L) | 364 | 1.2 (0.6) | 267 | 1.2 (0.6) |  | 0.360 |

Values are means (standard deviations) for continuous variables and n (%) for categorical/binary. For continuous variables, analyses were conducted with ANOVA and assumptions of normality inspected visually

## Discussion

Based on our data, the genetics of CRF may offer some protection against common NCDs, particularly T2D. However, no health discrepancies were observed between the “inherited” and “gained” CRF groups, which supports the importance of high CRF for health regardless of whether it is inherited or acquired through exercise.

Our study provides novel insights into how the genetics of CRF contribute to health. First, our analyses suggest that genetic contribution is approximately 2% risk reduction in T2D per SD increase in PGS CRF. For comparison, the risk for incident T2D is estimated to decrease by 8% per 1-MET increase in CRF.^6^ Overall, studies utilising genome-wide genetic instruments for CRF and health outcomes are limited, but existing data support our findings regarding T2D,^29^ longevity,^34^ and partially those related to CVD and cancer,^35,36^ Differences in methods (e.g. varying genetic instruments for CRF) and analytical approaches (mainly Mendelian randomisation) challenge the comparability of studies.

Second, previous studies reporting high heritability estimates for CRF (ranging from 44% to 68%)^8^ have utilised family^37^ or twin designs.^38^ The distinction between these study designs and ours is important when interpreting the results. Our study quantified genetics using measured and common variants (MAF >5%). This approach was chosen to evaluate the public health relevance of CRF genetics, i.e. associations expected to be commonly observed in the population. However, the findings do not diminish the significance of potential rare genetic variants which have larger effect sizes. While such variants may be highly relevant for individuals and families, their relevance in a public health context needs further study.

Third, more awareness and evidence are needed to address CRF as a complex, polygenic trait. In polygenic traits, single SNPs explain individually only a small proportion of a phenotype.^39^ Although computational genetics are constantly evolving, many scoring methods remain dependent on GWASs. Relevant GWASs are preferably based on large sample sizes to capture adequate genetic variation while including a well-phenotyped trait of interest in a population similar to the one with which the scores are to be used. The current GWASs for CRF may align well with the broader construct of physical fitness (i.e. ability to perform physical activity), but are insufficiently harmonised with respect to the definition of CRF (i.e. the body’s ability to deliver and utilise oxygen for work).^9,10,29^ However, since their initial releases in early 2020s, GWASs have brought new insights into the genetic basis of CRF.^7^ Based on current data, CRF genetics appear to be a mixture of SNPs with both positive and negative associations.^9,10,29^ Thus, CRF genetics may reflect not only favourable body structure and function but also absence of disease. For example, genes replicated in two UK Biobank-based studies, *CCDC141* and *KIAA1755*, have shown positive and negative associations with CRF, respectively.^9,29^ Of note, *CCDC141* has a role in the regulation of heart rate and blood pressure, while *KIAA1755* has been associated with an increased risk of atrial fibrillation, cardioembolic stroke, and metabolic syndrome.^40^ Our study supports these observations, especially for T2D but partially also for CVD, although adiposity may be an important factor in the causal framework. However, difficulties in replicating observed SNPs across different populations remain a challenge, and additional studies are needed to advance the field.^7^

### Research implications

Current GWASs for CRF range from performance phenotypes (watts/kg)^9^ to resting heart rate-derived metrics^29^ and estimated^29^ or directly measured oxygen consumption,^10^ utilising various scaling approaches (per body weight, per fat-free mass).^9,10,29^ In the future, a novel GWAS is recommended to be developed following a careful psychometric evaluation; what constitutes CRF, what does not, and which metrics demonstrate strong construct and criterion validity. More genetically informed studies using harmonised CRF testing are needed.

### Strengths and limitations

The study has several strengths, such as the use and evaluation of a novel genome-wide genetic score, the availability of gold standard measurement for CRF, and a robust replication design with two independent populations. The study is not without limitations, however. The current PGS CRF explains a small proportion of the variation in CRF, and the results need to be replicated when methods and available data have improved. Also, the PGS was constructed based on submaximal fitness test results, and although other PGSs did not show better validity, they might have different health associations. FinnGen has limited data on covariates, and thus, residual confounding may bias the findings. FinnGen is based mainly on individuals having an established contact with healthcare services,^16^ and this may limit the generalizability of the survival analyses.

## Conclusions

We found that a higher PGS CRF was associated with a lower risk of incident T2D, but not with cardiovascular disease, cancers, pulmonary disease, and all-cause mortality after controlling for potential covariates. Furthermore, as hypothesised, no differences in health characteristics and disease incidence were observed between “inherited” and “gained” CRF, supporting the importance of high CRF for health.

## Statements

### Contributorship

LJ and ES conceptualised the research question, and LJ wrote a statistical analysis plan to which all authors contributed. LJ preprocessed the publicly available genetic data. VL calculated the polygenic scores for FinnGen with the help of PH, and LJ calculated the scores for HUNT with the help of NPT. VL performed statistical modelling in FinnGen under the supervision of LJ, PH, MO, and ES. LJ performed the analyses in the HUNT in consultation with NPT, ANN, MK, KØ, and AB. ES secured access to FinnGen data.

FinnGen authors and their roles in data management and collection are presented in the Supplement. Additionally UK contributed to FinnGen data collection. ANN, MK, KØ, AB, and UW contributed to HUNT data collection and management. LJ, VL, and PH drafted the first version of the manuscript, and all authors contributed significantly to the writing, critical interpretation of the findings, and revising the manuscript. ES acquired funding for the study. LJ is the guarantor and attests that all listed authors meet authorship criteria and that no others meeting the criteria have been omitted.

### Competing interests

The authors declare no competing interests.

### Funding

The funders did not affect this study in any way. This work was funded by Research Council of Finland (grants 341750, 346509, and 361981), Juho Vainio Foundation; Päivikki and Sakari Sohlberg Foundation, all to ES. VL is funded by a University of Helsinki doctoral researcher position in the iCANDOC Doctoral Education Pilot in Precision Cancer Medicine. The FinnGen project is funded by Business Finland and 13 international pharmaceutical industry partners: AbbVie, AstraZeneca, Biogen, Boehringer Ingelheim, Celgene/Bristol-Myers Scibb, Genentech (a member of the Roche Group), GSK, Janssen, Maze Therapeutics, MSD/Merck, Novartis, Pfizer, and Sanofi. The HUNT study is a collaboration between HUNT Research Center (Faculty of Medicine and Health Sciences, Norwegian University of Science and Technology, NTNU), Trøndelag County Council, Central Norway Regional Health Authority, and the Norwegian Institute of Public Health. The genotyping in HUNT was financed by the National Institutes of Health; University of Michigan; the Research Council of Norway; the Liaison Committee for Education, Research and Innovation in Central Norway; and the Joint Research Committee between St Olavs Hospital and the Faculty of Medicine and Health Sciences, NTNU. The genetic investigations of the HUNT study are a collaboration between researchers from the K.G. Jebsen Center for Genetic Epidemiology, NTNU, and University of Michigan Medical School, and the University of Michigan School of Public Health. The K.G. Jebsen Center for Genetic Epidemiology is financed by Stiftelsen Kristian Gerhard Jebsen; Faculty of Medicine and Health Sciences, NTNU, Norway.

### Data sharing

Access to individual-level genotypes and register data from FinnGen participants can be applied for via the Fingenious portal (https://site.fingenious.fi/en/) hosted by the Finnish Biobank Cooperative FinBB (https://finbb.fi/en/).

Researchers affiliated with a Norwegian research institution can apply for HUNT data access from the HUNT Research Centre (www.ntnu.edu/hunt) if they have obtained project approval from the Regional Committee for Medical and Health Research Ethics (REC). Researchers not affiliated with a Norwegian research institution should collaborate with and apply through a Norwegian principal investigator. Information on the application and conditions for data access is available online (www.ntnu.edu/hunt/data).

### Ethical approval

Written informed consent was obtained from all participants, and the studies were approved by ethics committees. Participants in the FinnGen cohort provided informed consent for biobank research on the basis of the Finnish Biobank Act or cohort study-specific consent. The FinnGen study protocol (number HUS/990/2017) was approved by the Coordinating Ethics Committee of the Hospital District of Helsinki and Uusimaa. The FinnGen study is approved by the Finnish Institute of Health and Welfare (approval number THL/2031/6.02.00/2017), the Digital and Population Data Service Agency, the Social Insurance Institution, and Statistics Finland. The HUNT study was approved by the Regional Committee for Medical and Health Research Ethics (REC; 2019/29771), the Trøndelag Health Study, the Norwegian Data Inspectorate, and the National Directorate of Health. LJ is the guarantor and assumes responsibility for the accuracy and completeness of the protocol, data, and analyses, the fidelity of their reporting, and compliance with the Declaration of Helsinki, national laws, and the guidelines of the Finnish Advisory Board on Research Integrity.

### Transparency statement

LJ affirms that the manuscript is an honest, accurate, and transparent account of the study being reported; that no important aspects of the study have been omitted; and that any discrepancies from the study as originally planned (and, if relevant, registered) have been explained.

## Notes

### Competing Interest Statement

The authors have declared no competing interest.

### Author Declarations

Coordinating Ethics Committee of the Hospital District of Helsinki and Uusimaa approved the study the FinnGen study protocol (HUS/990/2017). The Finnish Institute of Health and Welfare, the Digital and Population Data Service Agency, the Social Insurance Institution, and Statistics Finland approved the FinnGen study (THL/2031/6.02.00/2017). The Regional Committee for Medical and Health Research Ethics, the Trøndelag Health Study, the Norwegian Data Inspectorate, and the National Directorate of Health approved the HUNT study (REC; 2019/29771).

