## Supplement for "Genetic Contribution of Cardiorespiratory Fitness in Morbidity and Mortality: A Prospective FinnGen and HUNT study"

Table S1. Endpoint data in FinnGen R12

| Phenotype | ICD-10 codes | FinnGen label | Cases/N |
|---|---|---|---|
| **Cardiovascular diseases** | | | |
| All CVDs | I00-I99 | I9_CVD | 284,261/ 520,210 |
| Ischemic heart diseases | I20-I25 | I9_ISCHHEART | 82,013 / 520,210 |
| Hypertension | I10-I15, I67.4 | I9_HYPTENS | 158,906 / 520,210 |
| Stroke | I60-I64, G45 | C_STROKE | 55,006 / 520,210 |
| **Cancers** | | | |
| All cancers | C00-C97 | ANY_CANC | 120,922 / 520,210 |
| Breast | C50 | C3_BREAST_EXALLC | 24,978 / 293,373 |
| Prostate | C61 | C3_PROSTATE_EXALLC | 20,966 / 226,837 |
| Colorectal | C18-20 | C3_COLORECTAL_EXALLC | 12187 / 520,210 |
| Lung# | C34 | C3_BRONCHUS_LUNG_EXALLC | 9,966 / 520,210 |
| **Pulmonary diseases** | | | |
| Chronic lower respiratory disease | J40-J47 | J10_LOWCHRON | 94,173 / 520,210 |
| Asthma | J45-46 | J10_ASTHMA | 61,122 / 520,210 |
| COPD | J43-44 | J10_COPD | 24,824 / 520,210 |
| **Metabolic diseases** | | | |
| Type 2 diabetes | E11* | E4_DM2 | 92,194 / 520,210 |

#, Bronchus and lung, *Additionally medicine purchases for blood glucose lowering drugs, excluding insulin. Endpoint data can be browsed using FinnGen labels in https://risteys.finregistry.fi/.

Table S2. Descriptives of individuals in HUNT3 with measured cardiorespiratory fitness

| Characteristics (N=3,655) | Mean (SD) or N (%) |
|---|---|
| Age (years) | 49.0 (13.1) |
| Sex, female n (%) | 1,920 (52.5%) |
| SES n (%) | |
| High | 1,674 (45.8%) |
| Medium | 1,825 (49.9%) |
| Low | 156 (4.3%) |
| BMI (kg/$m^2$) | 25.9 (3.5) |
| Smoking status n (%) | |
| Never | 1,830 (50.1%) |
| Former | 1,199 (32.8%) |
| Current | 626 (17.1%) |
| Self-reported health n (%) | |
| Poor | 9 (0.2%) |
| Not so good | 358 (9.8%) |
| Good | 2,229 (61.0%) |
| Very good | 1,059 (29.0%) |
| Physical activity (MET h/week) | 14.0 (9.1) |
| VO2max (ml/kg/min) | 40.3 (9.5) |
| VO2max (ml/$kg^{0.75}$/min) | 119.3 (28.4) |

A complete case sample of HUNT3 with genotype, VO2max and covariate data. Values are presented as means (standard deviation) for continuous variables and N (percentages) for others. SES, socioeconomic status; BMI, body mass index; MET, metabolic equivalent of task; VO2max, maximal oxygen consumption

Table S3. Associations between different genetic scores and selected measures of CRF.

| Genetic score | VO2max ml/kg/min | p | Adj.$R^2$ | VO2max ml/kg^0.75/min | p | Adj.$R^2$ |
|---|---|---|---|---|---|---|
| **PGS CRF** | 0.44 (0.25 to 0.63) | **<0.001** | 0.20% | 1.34 (0.79 to 1.89) | **<0.001** | 0.21% |
| **PGS CRF FFM** | 0.19 (0.01 to 0.38) | **0.040** | 0.03% | 0.58 (0.03 to 1.13) | **0.040** | 0.03% |
| **GRS CRF** | -0.03 (-0.22 to 0.16) | 0.775 | 0.00% | 0.01 (-0.54 to 0.56) | 0.975 | 0.00% |

Analyses were conducted using linear regression and interpreted as per one standard deviation increase in the genetic score. Values are unstandardized regression coefficients and 95% confidence intervals; All models were adjusted with first 10 principal components of ancestry, genotyping batch, age, sex, BMI, smoking status, socioeconomic status, self-reported health and self-reported physical activity (MET h/week); Adj.$R^2$, adjusted $R^2$ value for the genetic score in the model; PGS CRF, genome-wide polygenic score, constructed based on GWAS by Hanscombe et al. (2021) and the SBayesR method; PGS CRF FFM, genome-wide polygenic score constructed based on GWAS by Cai et al. (2023) and the SBayesR method; GRS CRF, genetic risk score, constructed based on favorable genetic variants reported by Cai et al. (2023) and their summarized effect sizes; Statistically significant associations highlighted with **bold**.

Table S4. A contingency table of the number of individuals in age- and sex-specific VO2max and PGS CRF deciles.

| VO2max decile | PGS CRF decile 10 | 9 | 8 | 7 | 6 | 5 | 4 | 3 | 2 | 1 |
|---|---|---|---|---|---|---|---|---|---|---|
| 10 | 58 | 41 | 37 | 31 | 33 | 26 | 37 | 35 | 28 | 29 |
| 9 | 42 | 38 | 49 | 34 | 35 | 34 | 33 | 28 | 28 | 34 |
| 8 | 36 | 45 | 29 | 38 | 37 | 42 | 34 | 29 | 43 | 25 |
| 7 | 38 | 31 | 31 | 34 | 35 | 33 | 32 | 35 | 42 | 50 |
| 6 | 39 | 42 | 35 | 37 | 43 | 36 | 30 | 40 | 32 | 29 |
| 5 | 27 | 32 | 28 | 31 | 38 | 42 | 43 | 47 | 32 | 46 |
| 4 | 33 | 41 | 44 | 40 | 38 | 34 | 35 | 35 | 33 | 35 |
| 3 | 34 | 32 | 38 | 42 | 36 | 38 | 41 | 37 | 44 | 31 |
| 2 | 30 | 35 | 33 | 35 | 38 | 45 | 41 | 42 | 37 | 41 |
| 1 | 28 | 28 | 41 | 43 | 32 | 36 | 40 | 38 | 47 | 46 |

Individuals with high VO2max and high PGS CRF are highlighted with blue ( , “inherited” CRF). Individuals with high VO2max and low PGS CRF are highlighted with green ( , “gained” CRF). Group sizes were based on a-priori sample size calculation.

### Table S5. Baseline characteristics of participants in the HUNT replication study

| Characteristics | All (n = 26,115) | Women (n = 13,687) | Men (n = 12,438) |
|---|---|---|---|
| Age (y) | 59.0 (14.5) | 58.6 (15.2) | 59.4 (13.4) |
| BMI ($kg/m^2$) | 27.8 (4.5) | 27.7 (5.0) | 27.9 (3.9) |
| Height (cm) | 170.1 (9.3) | 164.0 (6.3) | 177.0 (6.6) |
| Weight (kg) | 80.7 (15.4) | 74.4 (14.2) | 87.7 (13.6) |
| Smoking status n (%) | | | |
| Never | 9,902 (37.9) | 5,683 (41.5) | 4,219 (33.9) |
| Former | 9,611 (36.8) | 4,339 (31.7) | 5,272 (42.4) |
| Current | 6,602 (25.3) | 3,665 (26.8) | 2,937 (23.6) |

Values are means and (standard deviations) for continuous variables and n (%) for categorical/binary. BMI, body mass index.

### Table S6. Associations between polygenic score for cardiorespiratory fitness and morbidity in the HUNT cohort

| Disease | Cases/N | Model 1 HR (95% CI) | Model 2 HR (95% CI) | Model 3 HR (95% CI) |
|---|---|---|---|---|
| **Cardiovascular diseases** | 13,566/16,817 | 1.01 (0.99 to 1.03) | 1.01 (0.99 to 1.03) | 1.01 (0.99 to 1.03) |
| **Cancers** | 2,448/26,115 | 1.02 (0.99 to 1.07) | 1.02 (0.98 to 1.06) | 1.02 (0.98 to 1.06) |
| **Pulmonary diseases** | 3,185/25,084 | 0.97 (0.94 to 1.00) | 0.97 (0.94 to 1.01) | 0.98 (0.95 to 1.01) |
| **Type 2 diabetes** | 2,663/25,410 | **0.95 (0.92 to 0.99)** | 0.99 (0.95 to 1.03) | 0.99 (0.95 to 1.03) |

Values are hazard ratios (HR) and 95% confidence intervals (CI) per one standard deviation increase in polygenic score for cardiorespiratory fitness (PGS CRF). We used Cox proportional hazards models and age as the time scale. Model 1: PGS CRF, ten first principal components of ancestry, sex, and genotyping batch; Model 2, additionally body mass index; Model 3, additionally smoking status (never, former, current); Statistically significant associations highlighted with **bold**. No PGS CRF x Sex interactions were observed (all $p_{interactions}$ > 0.335) Mean follow-up was for cardiovascular diseases 8.1 years (min 0.1 years, max 18.1 years), for cancers 10.8 y (min 0.1 y, max 18.2 y), for pulmonary diseases 10.7 y (min 0.1, max 18.2 y) and for type 2 diabetes 10.7 y (min 0.1 y, max 18.2 y).

Table S7. Associations of “gained” cardiorespiratory fitness with morbidity in the HUNT cohort in comparison to “inherited” cardiorespiratory fitness

| Disease | Cases/N | HR (95% CI) | p-value |
|---|---|---|---|
| **Cardiovascular diseases** | 154/187 | | |
| “Inherited” CRF | 92/187 | Ref. | |
| “Gained” CRF | 62/187 | 0.83 (0.60 to 1.19) | 0.295 |
| **Cancers** | 34/231 | | |
| “Inherited” CRF | 20/231 | Ref. | |
| “Gained” CRF | 14/231 | 0.84 (0.41 to 1.69) | 0.626 |
| **Pulmonary diseases** | 12/232 | | |
| “Inherited” CRF | 8/232 | Ref. | |
| “Gained” CRF | 4/232 | 0.67 (0.20 to 2.24) | 0.515 |
| **Type 2 diabetes** | 3/231 | | |
| “Inherited” CRF | 2/231 | Ref. | |
| “Gained” CRF | 1/231 | 0.52 (0.05 to 5.90) | 0.594 |

Values are hazard ratios (HR) and 95% confidence intervals (CI) for belong to the “Gained” CRF group in comparison to “Inherited” CRF group. We used Cox proportional hazards models and age as the time scale. Model included the binary group variable but no covariates due to limited statistical power. Mean follow-up was for cardiovascular diseases 8.4 years (min 0.1 years, max 17.3 years), cancers 10.7 y (min 0.1 y, max 17.4 y), pulmonary diseases 11.1 y (min 0.1, max 17.4 y) and type 2 diabetes 11.0 y (min 0.1 y, max 17.4 y).

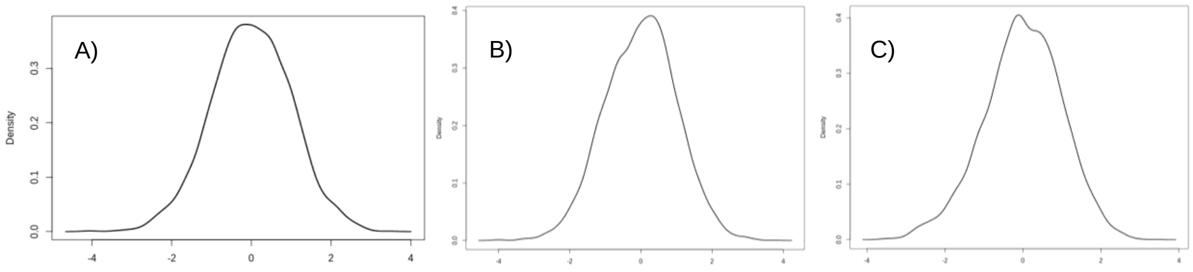


Figure S1. Density plots for three different genetic scores in the HUNT3; A) PGS CRF, B) PGS CRF FFM, and C) GRS CRF.

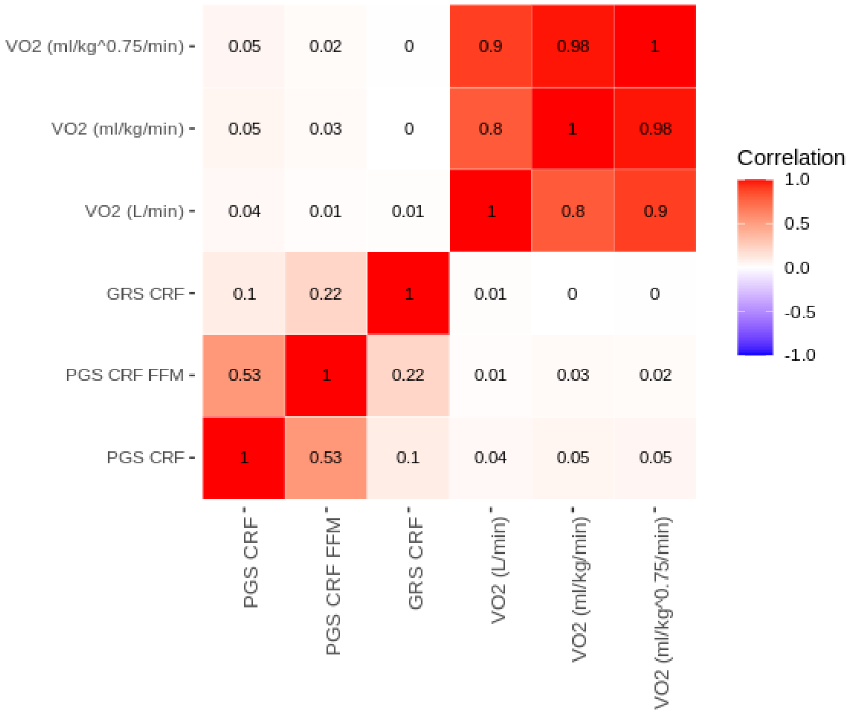


Figure S2. Correlation heatmap for PGS CRF, PGS CRF FFM and GRS CRF against selected markers of cardiorespiratory fitness. Values are Pearson correlation coefficients.

| Disease | Cases/N | Incidence/10,000 PY | HR (95% CI) | P-value |
|---|---|---|---|---|
| **Cardiovascular diseases** | | | | |
| Any CVD | 45,504 / 156,481 | 354.86 | 0.99 (0.98 to 1.00) | 0.120 |
| Ischemic heart diseases | 25,861 / 236,274 | 121.79 | 0.99 (0.97 to 1.00) | 0.029 |
| Hypertensive diseases | 31,533 / 208,759 | 171.60 | 1.00 (0.99 to 1.01) | 0.660 |
| Stroke | 18,247 / 199,684 | 79.70 | 0.99 (0.97 to 1.00) | 0.080 |
| **Cancer** | | | | |
| Any cancer | 29,570 / 227,855 | 138.59 | 1.00 (0.98 to 1.01) | 0.450 |
| Breast | 3,219 / 110,518 | 13.39 | 1.02 (0.98 to 1.05) | 0.360 |
| Prostate | 6,432 / 95,238 | 26.75 | 1.01 (0.98 to 1.03) | 0.670 |
| Colorectal | 3,230 / 199,335 | 13.23 | 0.98 (0.95 to 1.02) | 0.290 |
| Lung (Bronchus and lung) | 5,427 / 201,532 | 22.12 | 0.94 (0.92 to 0.97) | <0.001 |
| **Pulmonary diseases** | | | | |
| Chronic lower respiratory disease | 16,871 / 228,716 | 78.97 | 0.98 (0.97 to 1.00) | 0.017 |
| Asthma | 9,053 / 147,245 | 39.90 | 0.99 (0.97 to 1.01) | 0.230 |
| COPD | 10,553 / 222,398 | 44.35 | 0.97 (0.95 to 0.99) | 0.001 |
| **Metabolic diseases** | | | | |
| Type 2 diabetes | 21,347 / 233,842 | 99.71 | 0.97 (0.96 to 0.99) | <0.001 |
| **Mortality** | | | | |
| All-cause | 45,036 / 262,048 | 182.50 | 0.98 (0.97 to 0.99) | <0.001 |

0.9 0.95 1 1.05 1.1

← Lower risk Higher risk →

Figure S3. Associations of the PGS CRF with common NCDs and all-cause mortality following adjustment for BMI. The model was adjusted for sex, first 10 principal components of ancestry, genotyping batch and BMI at baseline.

| Disease | Cases/N | HR (95% CI) | P-value |
|---|---|---|---|
| **Cardiovascular diseases** | | | |
| Any CVD | 18,045 / 78,460 | 0.99 (0.98 to 1.00) | 0.17 |
| Ischemic heart diseases | 8,662 / 119,262 | 0.99 (0.97 to 1.01) | 0.44 |
| Hypertensive diseases | 13,170 / 103,988 | 1.00 (0.98 to 1.02) | 0.98 |
| Stroke | 6,910 / 104,939 | 1.00 (0.97 to 1.02) | 0.8 |
| **Cancer** | | | |
| Any cancer | 10,357 / 111,771 | 1.01 (0.99 to 1.03) | 0.2 |
| Breast | 2,005 / 66,448 | 1.01 (0.97 to 1.06) | 0.64 |
| Prostate | 1,726 / 37,588 | 1.02 (0.98 to 1.07) | 0.35 |
| Colorectal | 1,246 / 101,551 | 0.98 (0.92 to 1.03) | 0.38 |
| Lung (Bronchus and lung) | 443 / 100,748 | 1.01 (0.92 to 1.11) | 0.76 |
| **Pulmonary diseases** | | | |
| Chronic lower respiratory disease | 4,063 / 115,209 | 1.00 (0.96 to 1.03) | 0.77 |
| Asthma | 3,471 / 78,970 | 1.00 (0.96 to 1.03) | 0.79 |
| COPD | 509 / 111,655 | 0.96 (0.88 to 1.05) | 0.4 |
| **Metabolic diseases** | | | |
| Type 2 diabetes | 7,974 / 117,275 | 0.98 (0.95 to 1.00) | 0.028 |
| **Mortality** | | | |
| All-cause | 12,183 / 130,159 | 1.00 (0.98 to 1.02) | 0.77 |

0.9 0.95 1 1.05 1.1

← Lower risk Higher risk →

Figure S4. Associations of the PGS CRF with common NCDs and all-cause mortality among never-smokers. The model was adjusted for sex, first 10 principal components of ancestry, genotyping batch and BMI at baseline.

FinnGen Authors

| Full Name | Affiliation | Role 1 | Role 2 |
|---|---|---|---|
| Aarno Palotie | Institute for Molecular Medicine Finland (FIMM), HiLIFE, University of Helsinki, Helsinki, Finland; Broad Institute of MIT and Harvard; Massachusetts General Hospital, Boston, MA, United States | Steering Committee | Steering Committee |
| Mark Daly | Institute for Molecular Medicine Finland (FIMM), HiLIFE, University of Helsinki, Helsinki, Finland; Broad Institute of MIT and Harvard; Massachusetts General Hospital, Boston, MA, United States | Steering Committee | Steering Committee |
| Bridget Riley-Gills | Abbvie, Chicago, IL, United States | Steering Committee | Pharmaceutical companies |
| Howard Jacob | Abbvie, Chicago, IL, United States | Steering Committee | Pharmaceutical companies |
| Coralie Viollet | Astra Zeneca, Cambridge, United Kingdom | Steering Committee | Pharmaceutical companies |
| Slavé Petrovski | Astra Zeneca, Cambridge, United Kingdom | Steering Committee | Pharmaceutical companies |
| Alix Berton | Bayer AG, Leverkusen, Germany | Steering Committee | Pharmaceutical companies |
| Santha Ramakrishnan | Bayer AG, Leverkusen, Germany | Steering Committee | Pharmaceutical companies |
| Ellen Tsai | Biogen, Cambridge, MA, United States | Steering Committee | Pharmaceutical companies |
| Zhihao Ding | Boehringer Ingelheim, Ingelheim am Rhein, Germany | Steering Committee | Pharmaceutical companies |
| Emily Holzinger | Bristol Myers Squibb, New York, NY, United States | Steering Committee | Pharmaceutical companies |
| Robert Plenge | Bristol Myers Squibb, New York, NY, United States | Steering Committee | Pharmaceutical companies |
| Joseph Maranville | Bristol Myers Squibb, New York, NY, United States | Steering Committee | Pharmaceutical companies |
| Mark McCarthy | Genentech, San Francisco, CA, United States | Steering Committee | Pharmaceutical companies |
| Rion Pendergrass | Genentech, San Francisco, CA, United States | Steering Committee | Pharmaceutical companies |
| Jonathan Davitte | GlaxoSmithKline, Collegeville, PA, United States | Steering Committee | Pharmaceutical companies |
| Simonne Longerich | Merck, Kenilworth, NJ, United States | Steering Committee | Pharmaceutical companies |
| Anders Mälarstig | Pfizer, New York, NY, United States | Steering Committee | Pharmaceutical companies |
| Anna Vlahiotis | Pfizer, New York, NY, United States | Steering Committee | Pharmaceutical companies |
| Katherine Klinger | Translational Sciences, Sanofi R&D, Framingham, MA, USA | Steering Committee | Pharmaceutical companies |
| Clement Chatelain | Translational Sciences, Sanofi R&D, Framingham, MA, USA | Steering Committee | Pharmaceutical companies |
| Jorg Blankenstein | Translational Sciences, Sanofi R&D, Framingham, MA, USA | Steering Committee | Pharmaceutical companies |
| Karol Estrada | Maze Therapeutics, San Francisco, CA, United States | Steering Committee | Pharmaceutical companies |
| Robert Graham | Maze Therapeutics, San Francisco, CA, United States | Steering Committee | Pharmaceutical companies |
| Dawn Waterworth | Johnson & Johnson Innovative Medicine, Spring House, PA, United States | Steering Committee | Pharmaceutical companies |
| Chris O´Donnell | Novartis Institutes for BioMedical Research, Cambridge, MA, United States | Steering Committee | Pharmaceutical companies |
| Nicole Renaud | Novartis Institutes for BioMedical Research, Cambridge, MA, United States | Steering Committee | Pharmaceutical companies |
| Tomi P. Mäkelä | HiLIFE, University of Helsinki, Finland | Steering Committee | University of Helsinki & Biobanks |
| Jaakko Kaprio | Institute for Molecular Medicine Finland (FIMM), HiLIFE, University of Helsinki, Helsinki, Finland | Steering Committee | University of Helsinki & Biobanks |
| Minna Ruddock | Arctic biobank / University of Oulu, Oulu, Finland | Steering Committee | University of Helsinki & Biobanks |
| Petri Virolainen | Auria Biobank / University of Turku / Wellbeing Services County of Southwest Finland, Turku, Finland | Steering Committee | University of Helsinki & Biobanks |
| Antti Hakanen | Auria Biobank / University of Turku / Wellbeing Services County of Southwest Finland, Turku, Finland | Steering Committee | University of Helsinki & Biobanks |
| Terhi Kilpi | THL Biobank / Finnish Institute for Health and Welfare (THL), Helsinki, Finland | Steering Committee | University of Helsinki & Biobanks |
| Markus Perola | THL Biobank / Finnish Institute for Health and Welfare (THL), Helsinki, Finland | Steering Committee | University of Helsinki & Biobanks |

| | | | |
|---|---|---|---|
| Jukka Partanen | Finnish Red Cross Blood Service / Finnish Hematology Registry and Clinical Biobank, Helsinki, Finland | Steering Committee | **University of Helsinki & Biobanks** |
| Taneli Raivio | Helsinki Biobank / Helsinki University and Hospital District of Helsinki and Uusimaa, Helsinki | Steering Committee | **University of Helsinki & Biobanks** |
| Raisa Serpi | Northern Finland Biobank Borealis / University of Oulu / Wellbeing services county of North Ostrobothnia, Oulu, Finland | Steering Committee | **University of Helsinki & Biobanks** |
| Teija Kekonen | Northern Finland Biobank Borealis / University of Oulu / Wellbeing services county of North Ostrobothnia, Oulu, Finland | Steering Committee | **University of Helsinki & Biobanks** |
| Kati Kristiansson | Finnish Clinical Biobank Tampere / University of Tampere / Wellbeing Services County of Pirkanmaa, Tampere, Finland | Steering Committee | **University of Helsinki & Biobanks** |
| Veli-Matti Kosma | Biobank of Eastern Finland / University of Eastern Finland / Wellbeing services county of North Savo, Kuopio, Finland | Steering Committee | **University of Helsinki & Biobanks** |
| Jari Laukkanen | Central Finland Biobank / University of Jyväskylä / Wellbeing Services County of Central Finland, Jyväskylä, Finland | Steering Committee | **University of Helsinki & Biobanks** |
| Tom Southerington | Finnish Biobank Cooperative - FINBB | Steering Committee | **University of Helsinki & Biobanks** |
| Outi Tuovila | Business Finland, Helsinki, Finland | Steering Committee | **Other Experts/ Non-Voting Members** |
| Jeffrey Waring | Abbvie, Chicago, IL, United States | Scientific Committee | **Pharmaceutical companies** |
| Bridget Riley-Gillis | Abbvie, Chicago, IL, United States | Scientific Committee | **Pharmaceutical companies** |
| Fedik Rahimov | Abbvie, Chicago, IL, United States | Scientific Committee | **Pharmaceutical companies** |
| Ioanna Tachmazidou | Astra Zeneca, Cambridge, United Kingdom | Scientific Committee | **Pharmaceutical companies** |
| Alix Berton | Bayer AG, Leverkusen, Germany | Scientific Committee | **Pharmaceutical companies** |
| Santha Ramakrishnan | Bayer AG, Leverkusen, Germany | Scientific Committee | **Pharmaceutical companies** |
| Ellen Tsai | Biogen, Cambridge, MA, United States | Scientific Committee | **Pharmaceutical companies** |
| Zhihao Ding | Boehringer Ingelheim, Ingelheim am Rhein, Germany | Scientific Committee | **Pharmaceutical companies** |
| Marc Jung | Boehringer Ingelheim, Ingelheim am Rhein, Germany | Scientific Committee | **Pharmaceutical companies** |
| Hanati Tuoken | Boehringer Ingelheim, Ingelheim am Rhein, Germany | Scientific Committee | **Pharmaceutical companies** |
| Shameek Biswas | Bristol Myers Squibb, New York, NY, United States | Scientific Committee | **Pharmaceutical companies** |
| Benjamin Sun | Bristol Myers Squibb, New York, NY, United States | Scientific Committee | **Pharmaceutical companies** |
| Rion Pendergrass | Genentech, San Francisco, CA, United States | Scientific Committee | **Pharmaceutical companies** |
| Jonathan Davitte | GlaxoSmithKline, Collegeville, PA, United States | Scientific Committee | **Pharmaceutical companies** |
| Neha Raghavan | Merck, Kenilworth, NJ, United States | Scientific Committee | **Pharmaceutical companies** |
| Adriana Huertas-Vazquez | Merck, Kenilworth, NJ, United States | Scientific Committee | **Pharmaceutical companies** |
| Jae-Hoon Sul | Merck, Kenilworth, NJ, United States | Scientific Committee | **Pharmaceutical companies** |
| Anders Mälarstig | Pfizer, New York, NY, United States | Scientific Committee | **Pharmaceutical companies** |
| Xinli Hu | Pfizer, New York, NY, United States | Scientific Committee | **Pharmaceutical companies** |
| Åsa Hedman | Pfizer, New York, NY, United States | Scientific Committee | **Pharmaceutical companies** |
| Katherine Klinger | Translational Sciences, Sanofi R&D, Framingham, MA, USA | Scientific Committee | **Pharmaceutical companies** |
| Robert Graham | Maze Therapeutics, San Francisco, CA, United States | Scientific Committee | **Pharmaceutical companies** |
| Dawn Waterworth | Johnson & Johnson Innovative Medicine, Spring House, PA, United States | Scientific Committee | **Pharmaceutical companies** |
| Nicole Renaud | Novartis Institutes for BioMedical Research, Cambridge, MA, United States | Scientific Committee | **Pharmaceutical companies** |
| Ma´en Obeidat | Novartis Institutes for BioMedical Research, Cambridge, MA, United States | Scientific Committee | **Pharmaceutical companies** |
| Jonathan Chung | Novartis Institutes for BioMedical Research, Cambridge, MA, United States | Scientific Committee | **Pharmaceutical companies** |

| | | | |
|---|---|---|---|
| Jonas Zierer | Novartis Institutes for BioMedical Research, Cambridge, MA, United States | **Scientific Committee** | **Pharmaceutical companies** |
| Mari Niemi | Novartis Institutes for BioMedical Research, Cambridge, MA, United States | **Scientific Committee** | **Pharmaceutical companies** |
| Samuli Ripatti | Institute for Molecular Medicine Finland (FIMM), HiLIFE, University of Helsinki, Helsinki, Finland | **Scientific Committee** | **University of Helsinki & Biobanks** |
| Johanna Schleutker | Auria Biobank / University of Turku / Wellbeing Services County of Southwest Finland, Turku, Finland | **Scientific Committee** | **University of Helsinki & Biobanks** |
| Markus Perola | THL Biobank / Finnish Institute for Health and Welfare (THL), Helsinki, Finland | **Scientific Committee** | **University of Helsinki & Biobanks** |
| Mikko Arvas | Finnish Red Cross Blood Service / Finnish Hematology Registry and Clinical Biobank, Helsinki, Finland | **Scientific Committee** | **University of Helsinki & Biobanks** |
| Olli Carpén | Helsinki Biobank / Helsinki University and Hospital District of Helsinki and Uusimaa, Helsinki | **Scientific Committee** | **University of Helsinki & Biobanks** |
| Reetta Hinttala | Northern Finland Biobank Borealis / University of Oulu / Wellbeing services county of North Ostrobothnia, Oulu, Finland | **Scientific Committee** | **University of Helsinki & Biobanks** |
| Johannes Kettunen | Northern Finland Biobank Borealis / University of Oulu / Wellbeing services county of North Ostrobothnia, Oulu, Finland | **Scientific Committee** | **University of Helsinki & Biobanks** |
| Arto Mannermaa | Biobank of Eastern Finland / University of Eastern Finland / Wellbeing services county of North Savo, Kuopio, Finland | **Scientific Committee** | **University of Helsinki & Biobanks** |
| Katriina Aalto-Setälä | Faculty of Medicine and Health Technology, Tampere University, Tampere, Finland | **Scientific Committee** | **University of Helsinki & Biobanks** |
| Mika Kähönen | Finnish Clinical Biobank Tampere / University of Tampere / Wellbeing Services County of Pirkanmaa, Tampere, Finland | **Scientific Committee** | **University of Helsinki & Biobanks** |
| Jari Laukkanen | Central Finland Biobank / University of Jyväskylä / Wellbeing Services County of Central Finland, Jyväskylä, Finland | **Scientific Committee** | **University of Helsinki & Biobanks** |
| Johanna Mäkelä | FINBB - Finnish biobank cooperative | **Scientific Committee** | **University of Helsinki & Biobanks** |
| Lila Kallio | Auria Biobank / University of Turku / Wellbeing Services County of Southwest Finland, Turku, Finland | **Biobank directors** | **Biobank directors** |
| Tiina Wahlfors | THL Biobank / Finnish Institute for Health and Welfare (THL), Helsinki, Finland | **Biobank directors** | **Biobank directors** |
| Jukka Partanen | Finnish Red Cross Blood Service / Finnish Hematology Registry and Clinical Biobank, Helsinki, Finland | **Biobank directors** | **Biobank directors** |
| Eero Punkka | Helsinki Biobank / Helsinki University and Hospital District of Helsinki and Uusimaa, Helsinki | **Biobank directors** | **Biobank directors** |
| Raisa Serpi | Northern Finland Biobank Borealis / University of Oulu / Wellbeing services county of North Ostrobothnia, Oulu, Finland | **Biobank directors** | **Biobank directors** |
| Sanna Siltanen | Finnish Clinical Biobank Tampere / University of Tampere / Wellbeing Services County of Pirkanmaa, Tampere, Finland | **Biobank directors** | **Biobank directors** |
| Veli-Matti Kosma | Biobank of Eastern Finland / University of Eastern Finland / Wellbeing services county of North Savo, Kuopio, Finland | **Biobank directors** | **Biobank directors** |
| Tiina Jokela | Central Finland Biobank / University of Jyväskylä / Wellbeing Services County of Central Finland, Jyväskylä, Finland | **Biobank directors** | **Biobank directors** |
| Anu Jalanko | Institute for Molecular Medicine Finland (FIMM), HiLIFE, University of Helsinki, Helsinki, Finland | **FinnGen Teams** | **Administration** |
| Auli Toivola | Institute for Molecular Medicine Finland (FIMM), HiLIFE, University of Helsinki, Helsinki, Finland | **FinnGen Teams** | **Administration** |
| Denise Öller | Institute for Molecular Medicine Finland (FIMM), HiLIFE, University of Helsinki, Helsinki, Finland | **FinnGen Teams** | **Administration** |
| Helen Cooper | Institute for Molecular Medicine Finland (FIMM), HiLIFE, University of Helsinki, Helsinki, Finland | **FinnGen Teams** | **Administration** |
| Mervi Aavikko | Institute for Molecular Medicine Finland (FIMM), HiLIFE, University of Helsinki, Helsinki, Finland | **FinnGen Teams** | **Administration** |
| Risto Kajanne | Institute for Molecular Medicine Finland (FIMM), HiLIFE, University of Helsinki, Helsinki, Finland | **FinnGen Teams** | **Administration** |
| Rodos Rodosthenous | Institute for Molecular Medicine Finland (FIMM), HiLIFE, University of Helsinki, Helsinki, Finland | **FinnGen Teams** | **Administration** |
| Sofia Kuitunen | University of Helsinki, Helsinki, Finland | **FinnGen Teams** | **Administration** |
| Tarja Laitinen | Institute for Molecular Medicine Finland (FIMM), HiLIFE, University of Helsinki, Helsinki, Finland | **FinnGen Teams** | **Administration** |

| | | | |
|---|---|---|---|
| Arto Lehisto | Institute for Molecular Medicine Finland (FIMM), HiLIFE, University of Helsinki, Helsinki, Finland | FinnGen Teams | **Analysis** |
| Hafiz Sikandar | Institute for Molecular Medicine Finland (FIMM), HiLIFE, University of Helsinki, Helsinki, Finland | FinnGen Teams | **Analysis** |
| Juha Karjalainen | Institute for Molecular Medicine Finland (FIMM), HiLIFE, University of Helsinki, Helsinki, Finland | FinnGen Teams | **Analysis** |
| Juha Mehtonen | Institute for Molecular Medicine Finland (FIMM), HiLIFE, University of Helsinki, Helsinki, Finland | FinnGen Teams | **Analysis** |
| Masahiro Kanai | Broad Institute, Cambridge, MA, United States | FinnGen Teams | **Analysis** |
| Mitja Kurki | Institute for Molecular Medicine Finland (FIMM), HiLIFE, University of Helsinki, Helsinki, Finland; Broad Institute, Cambridge, MA, United States | FinnGen Teams | **Analysis** |
| Mutaamba Maasha | Broad Institute, Cambridge, MA, United States | FinnGen Teams | **Analysis** |
| Pietro Della Briotta Parolo | Institute for Molecular Medicine Finland (FIMM), HiLIFE, University of Helsinki, Helsinki, Finland | FinnGen Teams | **Analysis** |
| Samuel Jones | Institute for Molecular Medicine Finland (FIMM), HiLIFE, University of Helsinki, Helsinki, Finland | FinnGen Teams | **Analysis** |
| Sanni Ruotsalainen | Institute for Molecular Medicine Finland (FIMM), HiLIFE, University of Helsinki, Helsinki, Finland | FinnGen Teams | **Analysis** |
| Susanna Lemmelä | Institute for Molecular Medicine Finland (FIMM), HiLIFE, University of Helsinki, Helsinki, Finland | FinnGen Teams | **Analysis** |
| Wei Zhou | Broad Institute, Cambridge, MA, United States | FinnGen Teams | **Analysis** |
| Aki Havulinna | Finnish Institute for Health and Welfare (THL), Helsinki, Finland | FinnGen Teams | **Clinical Endpoint Development** |
| L. Elisa Lahtela | Institute for Molecular Medicine Finland (FIMM), HiLIFE, University of Helsinki, Helsinki, Finland | FinnGen Teams | **Clinical Endpoint Development** |
| Mari Kaunisto | Institute for Molecular Medicine Finland (FIMM), HiLIFE, University of Helsinki, Helsinki, Finland | FinnGen Teams | **Communication** |
| Awaisa Ghazal | Institute for Molecular Medicine Finland (FIMM), HiLIFE, University of Helsinki, Helsinki, Finland | FinnGen Teams | **E-Science** |
| Elina Kilpeläinen | Institute for Molecular Medicine Finland (FIMM), HiLIFE, University of Helsinki, Helsinki, Finland | FinnGen Teams | **E-Science** |
| Jaska Uimonen | Institute for Molecular Medicine Finland (FIMM), HiLIFE, University of Helsinki, Helsinki, Finland | FinnGen Teams | **E-Science** |
| Oluwaseun Alexander Dada | Institute for Molecular Medicine Finland (FIMM), HiLIFE, University of Helsinki, Helsinki, Finland | FinnGen Teams | **E-Science** |
| Rigbe Weldatsadik | Institute for Molecular Medicine Finland (FIMM), HiLIFE, University of Helsinki, Helsinki, Finland | FinnGen Teams | **E-Science** |
| Sanni Ruotsalainen | Institute for Molecular Medicine Finland (FIMM), HiLIFE, University of Helsinki, Helsinki, Finland | FinnGen Teams | **E-Science** |
| Tianduanyi Wang | Institute for Molecular Medicine Finland (FIMM), HiLIFE, University of Helsinki, Helsinki, Finland | FinnGen Teams | **E-Science** |
| Timo P. Sipilä | Institute for Molecular Medicine Finland (FIMM), HiLIFE, University of Helsinki, Helsinki, Finland | FinnGen Teams | **E-Science** |
| Kati Donner | Institute for Molecular Medicine Finland (FIMM), HiLIFE, University of Helsinki, Helsinki, Finland | FinnGen Teams | **Genotyping** |
| Anu Loukola | Helsinki Biobank / Helsinki University and Hospital District of Helsinki and Uusimaa, Helsinki | FinnGen Teams | **Sample Collection Coordination** |
| Päivi Ingalsuo | THL Biobank / Finnish Institute for Health and Welfare (THL), Helsinki, Finland | FinnGen Teams | **Sample Logistics** |
| Arto Pietilä | THL Biobank / Finnish Institute for Health and Welfare (THL), Helsinki, Finland | FinnGen Teams | **Registry Data Operations** |
| Sami Koskelainen | THL Biobank / Finnish Institute for Health and Welfare (THL), Helsinki, Finland | FinnGen Teams | **Registry Data Operations** |
| Susanna Lemmelä | Institute for Molecular Medicine Finland (FIMM), HiLIFE, University of Helsinki, Helsinki, Finland | FinnGen Teams | **Registry Data Operations** |
| Teemu Paajanen | THL Biobank / Finnish Institute for Health and Welfare (THL), Helsinki, Finland | FinnGen Teams | **Registry Data Operations** |
| Tero Hiekkalinna | THL Biobank / Finnish Institute for Health and Welfare (THL), Helsinki, Finland | FinnGen Teams | **Registry Data Operations** |
| Priit Palta | Institute for Molecular Medicine Finland (FIMM), HiLIFE, University of Helsinki, Helsinki, Finland | FinnGen Teams | **Sequencing Informatics** |
| Dawit A. Yohannes | Institute for Molecular Medicine Finland (FIMM), HiLIFE, University of Helsinki, Helsinki, Finland | FinnGen Teams | **Phenotype team** |
| Harri Siirtola | University of Tampere, Tampere, Finland | FinnGen Teams | **Phenotype team** |
| Javier Gracia-Tabuenca | University of Tampere, Tampere, Finland | FinnGen Teams | **Phenotype team** |
| Marika Kaakinen | Institute for Molecular Medicine Finland (FIMM), HiLIFE, University of Helsinki, Helsinki, Finland | FinnGen Teams | **Phenotype team** |
| Mary Pat Reeve | Institute for Molecular Medicine Finland (FIMM), HiLIFE, University of Helsinki, Helsinki, Finland | FinnGen Teams | **Phenotype team** |
| Shanmukha Sampath Padmanabhuni | Institute for Molecular Medicine Finland (FIMM), HiLIFE, University of Helsinki, Helsinki, Finland | FinnGen Teams | **Phenotype team** |

| | | | |
|---|---|---|---|
| Shuang Luo | Institute for Molecular Medicine Finland (FIMM), HiLIFE, University of Helsinki, Helsinki, Finland | FinnGen Teams | **Phenotype team** |
| Vincent Llorens | Institute for Molecular Medicine Finland (FIMM), HiLIFE, University of Helsinki, Helsinki, Finland | FinnGen Teams | **Phenotype team** |
| Iina Laak | Institute for Molecular Medicine Finland (FIMM), HiLIFE, University of Helsinki, Helsinki, Finland | FinnGen Teams | **Data protection officer** |
| Jaakko Tyrmi | University of Oulu, Oulu, Finland / University of Tampere, Tampere, Finland | FinnGen Teams | **FinnGen local support person** |
| Janne Isojärvi | University of Turku, Turku, Finland | FinnGen Teams | **FinnGen local support person** |
| Tero Sievänen | University of Eastern Finland, Kuopio, Finland | FinnGen Teams | **FinnGen local support person** |
| Timo Pohjonen | University of Jyväskylä, Jyväskylä, Finland | FinnGen Teams | **FinnGen local support person** |
| Vidal Fey | University of Tampere, Tampere, Finland | FinnGen Teams | **FinnGen local support person** |
| Johanna Mäkelä | Finnish Biobank Cooperative - FINBB | FinnGen Teams | **FINBB - Finnish biobank cooperative** |
| Pauli Wihuri | Finnish Biobank Cooperative - FINBB | FinnGen Teams | **FINBB - Finnish biobank cooperative** |
| Tom Southerington | Finnish Biobank Cooperative - FINBB | FinnGen Teams | **FINBB - Finnish biobank cooperative** |
| Meri Lähteenmäki | Finnish Biobank Cooperative - FINBB | FinnGen Teams | **FINBB - Finnish biobank cooperative** |
| Reetta Kälviäinen | Wellbeing services county of North Savo, Kuopio, Finland | Clinical Groups (FinnGen phases 1&2) | **Neurology Group** |
| Valtteri Julkunen | Wellbeing services county of North Savo, Kuopio, Finland | Clinical Groups (FinnGen phases 1&2) | **Neurology Group** |
| Hilkka Soininen | Wellbeing services county of North Savo, Kuopio, Finland | Clinical Groups (FinnGen phases 1&2) | **Neurology Group** |
| Anne Remes | Wellbeing services county of North Ostrobothnia, Oulu, Finland | Clinical Groups (FinnGen phases 1&2) | **Neurology Group** |
| Mikko Hiltunen | University of Eastern Finland, Kuopio, Finland | Clinical Groups (FinnGen phases 1&2) | **Neurology Group** |
| Jukka Peltola | Wellbeing Services County of Pirkanmaa, Tampere, Finland | Clinical Groups (FinnGen phases 1&2) | **Neurology Group** |
| Minna Raivio | Hospital District of Helsinki and Uusimaa, Helsinki, Finland | Clinical Groups (FinnGen phases 1&2) | **Neurology Group** |
| Pentti Tienari | Hospital District of Helsinki and Uusimaa, Helsinki, Finland | Clinical Groups (FinnGen phases 1&2) | **Neurology Group** |
| Juha Rinne | Wellbeing Services County of Southwest Finland, Turku, Finland | Clinical Groups (FinnGen phases 1&2) | **Neurology Group** |
| Roosa Kallionpää | Wellbeing Services County of Southwest Finland, Turku, Finland | Clinical Groups (FinnGen phases 1&2) | **Neurology Group** |
| Juulia Partanen | Institute for Molecular Medicine Finland, HiLIFE, University of Helsinki, Finland | Clinical Groups (FinnGen phases 1&2) | **Neurology Group** |
| Adam Ziemann | Abbvie, Chicago, IL, United States | Clinical Groups (FinnGen phases 1&2) | **Neurology Group** |
| Nizar Smaoui | Abbvie, Chicago, IL, United States | Clinical Groups (FinnGen phases 1&2) | **Neurology Group** |
| Anne Lehtonen | Abbvie, Chicago, IL, United States | Clinical Groups (FinnGen phases 1&2) | **Neurology Group** |
| Susan Eaton | Biogen, Cambridge, MA, United States | Clinical Groups (FinnGen phases 1&2) | **Neurology Group** |

| | | | |
|---|---|---|---|
| Shameek Biswas | Bristol Myers Squibb, New York, NY, United States | Clinical Groups (FinnGen phases 1&2) | **Neurology Group** |
| Natalie Bowers | Genentech, San Francisco, CA, United States | Clinical Groups (FinnGen phases 1&2) | **Neurology Group** |
| Edmond Teng | Genentech, San Francisco, CA, United States | Clinical Groups (FinnGen phases 1&2) | **Neurology Group** |
| Rion Pendergrass | Genentech, San Francisco, CA, United States | Clinical Groups (FinnGen phases 1&2) | **Neurology Group** |
| Fanli Xu | GlaxoSmithKline, Brentford, United Kingdom | Clinical Groups (FinnGen phases 1&2) | **Neurology Group** |
| Laura Addis | GlaxoSmithKline, Brentford, United Kingdom | Clinical Groups (FinnGen phases 1&2) | **Neurology Group** |
| John Eicher | GlaxoSmithKline, Brentford, United Kingdom | Clinical Groups (FinnGen phases 1&2) | **Neurology Group** |
| Qingqin S Li | Johnson & Johnson Innovative Medicine, Titusville, NJ 08560, United States | Clinical Groups (FinnGen phases 1&2) | **Neurology Group** |
| Karen He | Johnson & Johnson Innovative Medicine, Spring House, PA, United States | Clinical Groups (FinnGen phases 1&2) | **Neurology Group** |
| Ekaterina Khramtsova | Johnson & Johnson Innovative Medicine, Spring House, PA, United States | Clinical Groups (FinnGen phases 1&2) | **Neurology Group** |
| Neha Raghavan | Merck, Kenilworth, NJ, United States | Clinical Groups (FinnGen phases 1&2) | **Neurology Group** |
| Martti Färkkilä | Hospital District of Helsinki and Uusimaa, Helsinki, Finland | Clinical Groups (FinnGen phases 1&2) | **Gastroenterology Group** |
| Jukka Koskela | Hospital District of Helsinki and Uusimaa, Helsinki, Finland | Clinical Groups (FinnGen phases 1&2) | **Gastroenterology Group** |
| Sampsa Pikkarainen | Hospital District of Helsinki and Uusimaa, Helsinki, Finland | Clinical Groups (FinnGen phases 1&2) | **Gastroenterology Group** |
| Airi Jussila | Wellbeing Services County of Pirkanmaa, Tampere, Finland | Clinical Groups (FinnGen phases 1&2) | **Gastroenterology Group** |
| Katri Kaukinen | Wellbeing Services County of Pirkanmaa, Tampere, Finland | Clinical Groups (FinnGen phases 1&2) | **Gastroenterology Group** |
| Timo Blomster | Wellbeing services county of North Ostrobothnia, Oulu, Finland | Clinical Groups (FinnGen phases 1&2) | **Gastroenterology Group** |
| Mikko Kiviniemi | Wellbeing services county of North Savo, Kuopio, Finland | Clinical Groups (FinnGen phases 1&2) | **Gastroenterology Group** |
| Markku Voutilainen | Wellbeing Services County of Southwest Finland, Turku, Finland | Clinical Groups (FinnGen phases 1&2) | **Gastroenterology Group** |
| Mark Daly | Institute for Molecular Medicine Finland (FIMM), HiLIFE, University of Helsinki, Helsinki, Finland; Broad Institute of MIT and Harvard; Massachusetts General Hospital, Boston, MA, United States | Clinical Groups (FinnGen phases 1&2) | **Gastroenterology Group** |
| Jeffrey Waring | Abbvie, Chicago, IL, United States | Clinical Groups (FinnGen phases 1&2) | **Gastroenterology Group** |
| Nizar Smaoui | Abbvie, Chicago, IL, United States | Clinical Groups (FinnGen phases 1&2) | **Gastroenterology Group** |
| Fedik Rahimov | Abbvie, Chicago, IL, United States | Clinical Groups (FinnGen phases 1&2) | **Gastroenterology Group** |
| Anne Lehtonen | Abbvie, Chicago, IL, United States | Clinical Groups (FinnGen phases 1&2) | **Gastroenterology Group** |

| | | | |
|---|---|---|---|
| Tim Lu | Genentech, San Francisco, CA, United States | Clinical Groups (FinnGen phases 1&2) | **Gastroenterology Group** |
| Natalie Bowers | Genentech, San Francisco, CA, United States | Clinical Groups (FinnGen phases 1&2) | **Gastroenterology Group** |
| Rion Pendergrass | Genentech, San Francisco, CA, United States | Clinical Groups (FinnGen phases 1&2) | **Gastroenterology Group** |
| Linda McCarthy | GlaxoSmithKline, Brentford, United Kingdom | Clinical Groups (FinnGen phases 1&2) | **Gastroenterology Group** |
| Amy Hart | Johnson & Johnson Innovative Medicine, Spring House, PA, United States | Clinical Groups (FinnGen phases 1&2) | **Gastroenterology Group** |
| Meijian Guan | Johnson & Johnson Innovative Medicine, Spring House, PA, United States | Clinical Groups (FinnGen phases 1&2) | **Gastroenterology Group** |
| Jason Miller | Merck, Kenilworth, NJ, United States | Clinical Groups (FinnGen phases 1&2) | **Gastroenterology Group** |
| Kirsi Kalpala | Pfizer, New York, NY, United States | Clinical Groups (FinnGen phases 1&2) | **Gastroenterology Group** |
| Melissa Miller | Pfizer, New York, NY, United States | Clinical Groups (FinnGen phases 1&2) | **Gastroenterology Group** |
| Xinli Hu | Pfizer, New York, NY, United States | Clinical Groups (FinnGen phases 1&2) | **Gastroenterology Group** |
| Kari Eklund | Hospital District of Helsinki and Uusimaa, Helsinki, Finland | Clinical Groups (FinnGen phases 1&2) | **Rheumatology Group** |
| Antti Palomäki | Wellbeing Services County of Southwest Finland, Turku, Finland | Clinical Groups (FinnGen phases 1&2) | **Rheumatology Group** |
| Pia Isomäki | Wellbeing Services County of Pirkanmaa, Tampere, Finland | Clinical Groups (FinnGen phases 1&2) | **Rheumatology Group** |
| Laura Pirilä | Wellbeing Services County of Southwest Finland, Turku, Finland | Clinical Groups (FinnGen phases 1&2) | **Rheumatology Group** |
| Oili Kaipiainen-Seppänen | Wellbeing services county of North Savo, Kuopio, Finland | Clinical Groups (FinnGen phases 1&2) | **Rheumatology Group** |
| Johanna Huhtakangas | Wellbeing services county of North Ostrobothnia, Oulu, Finland | Clinical Groups (FinnGen phases 1&2) | **Rheumatology Group** |
| Nina Mars | Institute for Molecular Medicine Finland (FIMM), HiLIFE, University of Helsinki, Helsinki, Finland | Clinical Groups (FinnGen phases 1&2) | **Rheumatology Group** |
| Jeffrey Waring | Abbvie, Chicago, IL, United States | Clinical Groups (FinnGen phases 1&2) | **Rheumatology Group** |
| Fedik Rahimov | Abbvie, Chicago, IL, United States | Clinical Groups (FinnGen phases 1&2) | **Rheumatology Group** |
| Apinya Lertratanakul | Abbvie, Chicago, IL, United States | Clinical Groups (FinnGen phases 1&2) | **Rheumatology Group** |
| Nizar Smaoui | Abbvie, Chicago, IL, United States | Clinical Groups (FinnGen phases 1&2) | **Rheumatology Group** |
| Anne Lehtonen | Abbvie, Chicago, IL, United States | Clinical Groups (FinnGen phases 1&2) | **Rheumatology Group** |
| Coralie Viollet | AstraZeneca, Cambridge, United Kingdom | Clinical Groups (FinnGen phases 1&2) | **Rheumatology Group** |
| Marla Hochfeld | Bristol Myers Squibb, New York, NY, United States | Clinical Groups (FinnGen phases 1&2) | **Rheumatology Group** |

| | | | |
|---|---|---|---|
| Natalie Bowers | Genentech, San Francisco, CA, United States | Clinical Groups (FinnGen phases 1&2) | **Rheumatology Group** |
| Rion Pendergrass | Genentech, San Francisco, CA, United States | Clinical Groups (FinnGen phases 1&2) | **Rheumatology Group** |
| Jorge Esparza Gordillo | GlaxoSmithKline, Brentford, United Kingdom | Clinical Groups (FinnGen phases 1&2) | **Rheumatology Group** |
| Dawn Waterworth | Johnson & Johnson Innovative Medicine, Spring House, PA, United States | Clinical Groups (FinnGen phases 1&2) | **Rheumatology Group** |
| Fabiana Farias | Merck, Kenilworth, NJ, United States | Clinical Groups (FinnGen phases 1&2) | **Rheumatology Group** |
| Kirsi Kalpala | Pfizer, New York, NY, United States | Clinical Groups (FinnGen phases 1&2) | **Rheumatology Group** |
| Nan Bing | Pfizer, New York, NY, United States | Clinical Groups (FinnGen phases 1&2) | **Rheumatology Group** |
| Xinli Hu | Pfizer, New York, NY, United States | Clinical Groups (FinnGen phases 1&2) | **Rheumatology Group** |
| Tarja Laitinen | Wellbeing Services County of Pirkanmaa, Tampere, Finland | Clinical Groups (FinnGen phases 1&2) | **Pulmonology Group** |
| Margit Pelkonen | Wellbeing services county of North Savo, Kuopio, Finland | Clinical Groups (FinnGen phases 1&2) | **Pulmonology Group** |
| Paula Kauppi | Hospital District of Helsinki and Uusimaa, Helsinki, Finland | Clinical Groups (FinnGen phases 1&2) | **Pulmonology Group** |
| Hannu Kankaanranta | University of Gothenburg, Gothenburg, Sweden/ Seinäjoki Central Hospital, Seinäjoki, Finland/ Tampere University, Tampere, Finland | Clinical Groups (FinnGen phases 1&2) | **Pulmonology Group** |
| Terttu Harju | Wellbeing services county of North Ostrobothnia, Oulu, Finland | Clinical Groups (FinnGen phases 1&2) | **Pulmonology Group** |
| Riitta Lahesmaa | Wellbeing Services County of Southwest Finland, Turku, Finland | Clinical Groups (FinnGen phases 1&2) | **Pulmonology Group** |
| Nizar Smaoui | Abbvie, Chicago, IL, United States | Clinical Groups (FinnGen phases 1&2) | **Pulmonology Group** |
| Coralie Viollet | AstraZeneca, Cambridge, United Kingdom | Clinical Groups (FinnGen phases 1&2) | **Pulmonology Group** |
| Susan Eaton | Biogen, Cambridge, MA, United States | Clinical Groups (FinnGen phases 1&2) | **Pulmonology Group** |
| Hubert Chen | Genentech, San Francisco, CA, United States | Clinical Groups (FinnGen phases 1&2) | **Pulmonology Group** |
| Rion Pendergrass | Genentech, San Francisco, CA, United States | Clinical Groups (FinnGen phases 1&2) | **Pulmonology Group** |
| Natalie Bowers | Genentech, San Francisco, CA, United States | Clinical Groups (FinnGen phases 1&2) | **Pulmonology Group** |
| Joanna Betts | GlaxoSmithKline, Brentford, United Kingdom | Clinical Groups (FinnGen phases 1&2) | **Pulmonology Group** |
| Kirsi Auro | GlaxoSmithKline, Espoo, Finland | Clinical Groups (FinnGen phases 1&2) | **Pulmonology Group** |
| Rajashree Mishra | GlaxoSmithKline, Brentford, United Kingdom | Clinical Groups (FinnGen phases 1&2) | **Pulmonology Group** |
| Majd Mouded | Novartis, Basel, Switzerland | Clinical Groups (FinnGen phases 1&2) | **Pulmonology Group** |

| | | | |
|---|---|---|---|
| Debby Ngo | Novartis, Basel, Switzerland | Clinical Groups (FinnGen phases 1&2) | **Pulmonology Group** |
| Teemu Niiranen | University of Turku, Turku, Finland; Finnish Institute for Health and Welfare (THL), Helsinki, Finland | Clinical Groups (FinnGen phases 1&2) | **Cardiometabolic Diseases Group** |
| Felix Vaura | Finnish Institute for Health and Welfare (THL), Helsinki, Finland | Clinical Groups (FinnGen phases 1&2) | **Cardiometabolic Diseases Group** |
| Veikko Salomaa | Finnish Institute for Health and Welfare (THL), Helsinki, Finland | Clinical Groups (FinnGen phases 1&2) | **Cardiometabolic Diseases Group** |
| Kaj Metsärinne | Wellbeing Services County of Southwest Finland, Turku, Finland | Clinical Groups (FinnGen phases 1&2) | **Cardiometabolic Diseases Group** |
| Jenni Aittokallio | Wellbeing Services County of Southwest Finland, Turku, Finland | Clinical Groups (FinnGen phases 1&2) | **Cardiometabolic Diseases Group** |
| Mika Kähönen | Finnish Clinical Biobank Tampere / University of Tampere / Wellbeing Services County of Pirkanmaa, Tampere, Finland | Clinical Groups (FinnGen phases 1&2) | **Cardiometabolic Diseases Group** |
| Jussi Hernesniemi | Wellbeing Services County of Pirkanmaa, Tampere, Finland | Clinical Groups (FinnGen phases 1&2) | **Cardiometabolic Diseases Group** |
| Daniel Gordin | Hospital District of Helsinki and Uusimaa, Helsinki, Finland | Clinical Groups (FinnGen phases 1&2) | **Cardiometabolic Diseases Group** |
| Juha Sinisalo | Hospital District of Helsinki and Uusimaa, Helsinki, Finland | Clinical Groups (FinnGen phases 1&2) | **Cardiometabolic Diseases Group** |
| Marja-Riitta Taskinen | Hospital District of Helsinki and Uusimaa, Helsinki, Finland | Clinical Groups (FinnGen phases 1&2) | **Cardiometabolic Diseases Group** |
| Tiinamaija Tuomi | Institute for Molecular Medicine Finland (FIMM), HiLIFE, University of Helsinki, Helsinki, Finland; Hospital District of Helsinki and Uusimaa, Helsinki, Finland | Clinical Groups (FinnGen phases 1&2) | **Cardiometabolic Diseases Group** |
| Timo Hiltunen | Hospital District of Helsinki and Uusimaa, Helsinki, Finland | Clinical Groups (FinnGen phases 1&2) | **Cardiometabolic Diseases Group** |
| Jari Laukkanen | Central Finland Biobank / University of Jyväskylä / Wellbeing Services County of Central Finland, Jyväskylä, Finland | Clinical Groups (FinnGen phases 1&2) | **Cardiometabolic Diseases Group** |
| Amanda Elliott | Institute for Molecular Medicine Finland (FIMM), HiLIFE, University of Helsinki, Helsinki, Finland; Broad Institute, Cambridge, MA, USA and Massachusetts General Hospital, Boston, MA, USA | Clinical Groups (FinnGen phases 1&2) | **Cardiometabolic Diseases Group** |
| Mary Pat Reeve | Institute for Molecular Medicine Finland (FIMM), HiLIFE, University of Helsinki, Helsinki, Finland | Clinical Groups (FinnGen phases 1&2) | **Cardiometabolic Diseases Group** |
| Sanni Ruotsalainen | Institute for Molecular Medicine Finland (FIMM), HiLIFE, University of Helsinki, Helsinki, Finland | Clinical Groups (FinnGen phases 1&2) | **Cardiometabolic Diseases Group** |
| Dirk Paul | Astra Zeneca, Cambridge, United Kingdom | Clinical Groups (FinnGen phases 1&2) | **Cardiometabolic Diseases Group** |
| Natalie Bowers | Genentech, San Francisco, CA, United States | Clinical Groups (FinnGen phases 1&2) | **Cardiometabolic Diseases Group** |
| Rion Pendergrass | Genentech, San Francisco, CA, United States | Clinical Groups (FinnGen phases 1&2) | **Cardiometabolic Diseases Group** |
| Audrey Chu | GlaxoSmithKline, Brentford, United Kingdom | Clinical Groups (FinnGen phases 1&2) | **Cardiometabolic Diseases Group** |
| Dermot Reilly | Johnson & Johnson Innovative Medicine, Boston, MA, United States | Clinical Groups (FinnGen phases 1&2) | **Cardiometabolic Diseases Group** |
| Mike Mendelson | Novartis, Boston, MA, United States | Clinical Groups (FinnGen phases 1&2) | **Cardiometabolic Diseases Group** |
| Jaakko Parkkinen | Pfizer, New York, NY, United States | Clinical Groups (FinnGen phases 1&2) | **Cardiometabolic Diseases Group** |

| | | | |
|---|---|---|---|
| Melissa Miller | Pfizer, New York, NY, United States | Clinical Groups (FinnGen phases 1&2) | **Cardiometabolic Diseases Group** |
| Tuomo Meretoja | Helsinki University Hospital and University of Helsinki, Helsinki, Finland | Clinical Groups (FinnGen phases 1&2) | **Oncology Group** |
| Heikki Joensuu | Helsinki University Hospital and University of Helsinki, Helsinki, Finland | Clinical Groups (FinnGen phases 1&2) | **Oncology Group** |
| Olli Carpén | Hospital District of Helsinki and Uusimaa, Helsinki, Finland | Clinical Groups (FinnGen phases 1&2) | **Oncology Group** |
| Johanna Mattson | Hospital District of Helsinki and Uusimaa, Helsinki, Finland | Clinical Groups (FinnGen phases 1&2) | **Oncology Group** |
| Eveliina Salminen | Hospital District of Helsinki and Uusimaa, Helsinki, Finland | Clinical Groups (FinnGen phases 1&2) | **Oncology Group** |
| Annika Auranen | Wellbeing Services County of Pirkanmaa, Tampere, Finland | Clinical Groups (FinnGen phases 1&2) | **Oncology Group** |
| Peeter Karihtala | Helsinki University Hospital and University of Helsinki, Helsinki, Finland | Clinical Groups (FinnGen phases 1&2) | **Oncology Group** |
| Päivi Auvinen | Wellbeing services county of North Savo, Kuopio, Finland | Clinical Groups (FinnGen phases 1&2) | **Oncology Group** |
| Klaus Elenius | Wellbeing Services County of Southwest Finland, Turku, Finland | Clinical Groups (FinnGen phases 1&2) | **Oncology Group** |
| Johanna Schleutker | Wellbeing Services County of Southwest Finland, Turku, Finland | Clinical Groups (FinnGen phases 1&2) | **Oncology Group** |
| Esa Pitkänen | Institute for Molecular Medicine Finland (FIMM), HiLIFE, University of Helsinki, Helsinki, Finland | Clinical Groups (FinnGen phases 1&2) | **Oncology Group** |
| Nina Mars | Institute for Molecular Medicine Finland (FIMM), HiLIFE, University of Helsinki, Helsinki, Finland | Clinical Groups (FinnGen phases 1&2) | **Oncology Group** |
| Mark Daly | Institute for Molecular Medicine Finland (FIMM), HiLIFE, University of Helsinki, Helsinki, Finland; Broad Institute of MIT and Harvard; Massachusetts General Hospital, Boston, MA, United States | Clinical Groups (FinnGen phases 1&2) | **Oncology Group** |
| Relja Popovic | Abbvie, Chicago, IL, United States | Clinical Groups (FinnGen phases 1&2) | **Oncology Group** |
| Jeffrey Waring | Abbvie, Chicago, IL, United States | Clinical Groups (FinnGen phases 1&2) | **Oncology Group** |
| Bridget Riley-Gillis | Abbvie, Chicago, IL, United States | Clinical Groups (FinnGen phases 1&2) | **Oncology Group** |
| Anne Lehtonen | Abbvie, Chicago, IL, United States | Clinical Groups (FinnGen phases 1&2) | **Oncology Group** |
| Margarete Fabre | AstraZeneca, Cambridge, United Kingdom | Clinical Groups (FinnGen phases 1&2) | **Oncology Group** |
| Jennifer Schutzman | Genentech, San Francisco, CA, United States | Clinical Groups (FinnGen phases 1&2) | **Oncology Group** |
| Natalie Bowers | Genentech, San Francisco, CA, United States | Clinical Groups (FinnGen phases 1&2) | **Oncology Group** |
| Rion Pendergrass | Genentech, San Francisco, CA, United States | Clinical Groups (FinnGen phases 1&2) | **Oncology Group** |
| Diptee Kulkarni | GlaxoSmithKline, Brentford, United Kingdom | Clinical Groups (FinnGen phases 1&2) | **Oncology Group** |
| Alessandro Porello | Johnson & Johnson Innovative Medicine, Spring House, PA, United States | Clinical Groups (FinnGen phases 1&2) | **Oncology Group** |

| | | | |
|---|---|---|---|
| Andrey Loboda | Merck, Kenilworth, NJ, United States | Clinical Groups (FinnGen phases 1&2) | **Oncology Group** |
| Stefan McDonough | Pfizer, New York, NY, United States | Clinical Groups (FinnGen phases 1&2) | **Oncology Group** |
| Kai Kaarniranta | Wellbeing services county of North Savo, Kuopio, Finland; University of Lodz, Lodz, Poland | Clinical Groups (FinnGen phases 1&2) | **Opthalmology Group** |
| Joni A Turunen | Helsinki University Hospital and University of Helsinki, Helsinki, Finland; Folkhälsan Research Center, Helsinki, Finland | Clinical Groups (FinnGen phases 1&2) | **Opthalmology Group** |
| Terhi Ollila | Hospital District of Helsinki and Uusimaa, Helsinki, Finland | Clinical Groups (FinnGen phases 1&2) | **Opthalmology Group** |
| Hannu Uusitalo | Wellbeing Services County of Pirkanmaa, Tampere, Finland | Clinical Groups (FinnGen phases 1&2) | **Opthalmology Group** |
| Juha Karjalainen | Institute for Molecular Medicine Finland (FIMM), HiLIFE, University of Helsinki, Helsinki, Finland | Clinical Groups (FinnGen phases 1&2) | **Opthalmology Group** |
| Esa Pitkänen | Institute for Molecular Medicine Finland (FIMM), HiLIFE, University of Helsinki, Helsinki, Finland | Clinical Groups (FinnGen phases 1&2) | **Opthalmology Group** |
| Mengzhen Liu | Abbvie, Chicago, IL, United States | Clinical Groups (FinnGen phases 1&2) | **Opthalmology Group** |
| Erich Strauss | Genentech, San Francisco, CA, United States | Clinical Groups (FinnGen phases 1&2) | **Opthalmology Group** |
| Natalie Bowers | Genentech, San Francisco, CA, United States | Clinical Groups (FinnGen phases 1&2) | **Opthalmology Group** |
| Hao Chen | Genentech, San Francisco, CA, United States | Clinical Groups (FinnGen phases 1&2) | **Opthalmology Group** |
| Rion Pendergrass | Genentech, San Francisco, CA, United States | Clinical Groups (FinnGen phases 1&2) | **Opthalmology Group** |
| Kaisa Tasanen | Wellbeing services county of North Ostrobothnia, Oulu, Finland | Clinical Groups (FinnGen phases 1&2) | **Dermatology Group** |
| Laura Huilaja | Wellbeing services county of North Ostrobothnia, Oulu, Finland | Clinical Groups (FinnGen phases 1&2) | **Dermatology Group** |
| Katariina Hannula-Jouppi | Hospital District of Helsinki and Uusimaa, Helsinki, Finland | Clinical Groups (FinnGen phases 1&2) | **Dermatology Group** |
| Teea Salmi | Wellbeing Services County of Pirkanmaa, Tampere, Finland | Clinical Groups (FinnGen phases 1&2) | **Dermatology Group** |
| Sirkku Peltonen | Wellbeing Services County of Southwest Finland, Turku, Finland | Clinical Groups (FinnGen phases 1&2) | **Dermatology Group** |
| Leena Koulu | Wellbeing Services County of Southwest Finland, Turku, Finland | Clinical Groups (FinnGen phases 1&2) | **Dermatology Group** |
| Nizar Smaoui | Abbvie, Chicago, IL, United States | Clinical Groups (FinnGen phases 1&2) | **Dermatology Group** |
| Fedik Rahimov | Abbvie, Chicago, IL, United States | Clinical Groups (FinnGen phases 1&2) | **Dermatology Group** |
| Anne Lehtonen | Abbvie, Chicago, IL, United States | Clinical Groups (FinnGen phases 1&2) | **Dermatology Group** |
| David Choy | Genentech, San Francisco, CA, United States | Clinical Groups (FinnGen phases 1&2) | **Dermatology Group** |
| Rion Pendergrass | Genentech, San Francisco, CA, United States | Clinical Groups (FinnGen phases 1&2) | **Dermatology Group** |

| | | | |
|---|---|---|---|
| Dawn Waterworth | Johnson & Johnson Innovative Medicine, Spring House, PA, United States | Clinical Groups (FinnGen phases 1&2) | **Dermatology Group** |
| Kirsi Kalpala | Pfizer, New York, NY, United States | Clinical Groups (FinnGen phases 1&2) | **Dermatology Group** |
| Ying Wu | Pfizer, New York, NY, United States | Clinical Groups (FinnGen phases 1&2) | **Dermatology Group** |
| Pirkko Pussinen | Hospital District of Helsinki and Uusimaa, Helsinki, Finland | Clinical Groups (FinnGen phases 1&2) | **Odontology Group** |
| Aino Salminen | Hospital District of Helsinki and Uusimaa, Helsinki, Finland | Clinical Groups (FinnGen phases 1&2) | **Odontology Group** |
| Tuula Salo | Hospital District of Helsinki and Uusimaa, Helsinki, Finland | Clinical Groups (FinnGen phases 1&2) | **Odontology Group** |
| David Rice | Hospital District of Helsinki and Uusimaa, Helsinki, Finland | Clinical Groups (FinnGen phases 1&2) | **Odontology Group** |
| Pekka Nieminen | Hospital District of Helsinki and Uusimaa, Helsinki, Finland | Clinical Groups (FinnGen phases 1&2) | **Odontology Group** |
| Ulla Palotie | Hospital District of Helsinki and Uusimaa, Helsinki, Finland | Clinical Groups (FinnGen phases 1&2) | **Odontology Group** |
| Maria Siponen | Wellbeing services county of North Savo, Kuopio, Finland | Clinical Groups (FinnGen phases 1&2) | **Odontology Group** |
| Liisa Suominen | Wellbeing services county of North Savo, Kuopio, Finland | Clinical Groups (FinnGen phases 1&2) | **Odontology Group** |
| Päivi Mäntylä | Wellbeing services county of North Savo, Kuopio, Finland | Clinical Groups (FinnGen phases 1&2) | **Odontology Group** |
| Ulvi Gursoy | Wellbeing Services County of Southwest Finland, Turku, Finland | Clinical Groups (FinnGen phases 1&2) | **Odontology Group** |
| Vuokko Anttonen | Wellbeing services county of North Ostrobothnia, Oulu, Finland | Clinical Groups (FinnGen phases 1&2) | **Odontology Group** |
| Kirsi Sipilä | Oulu University Hospital and University of Oulu, Oulu, Finland | Clinical Groups (FinnGen phases 1&2) | **Odontology Group** |
| Rion Pendergrass | Genentech, San Francisco, CA, United States | Clinical Groups (FinnGen phases 1&2) | **Odontology Group** |
| Hannele Laivuori | Institute for Molecular Medicine Finland (FIMM), HiLIFE, University of Helsinki, Helsinki, Finland | Clinical Groups (FinnGen phases 1&2) | **Women's Health and Reproduction Group** |
| Venla Kurra | Wellbeing Services County of Pirkanmaa, Tampere, Finland | Clinical Groups (FinnGen phases 1&2) | **Women's Health and Reproduction Group** |
| Laura Kotaniemi-Talonen | Wellbeing Services County of Pirkanmaa, Tampere, Finland | Clinical Groups (FinnGen phases 1&2) | **Women's Health and Reproduction Group** |
| Oskari Heikinheimo | Hospital District of Helsinki and Uusimaa, Helsinki, Finland | Clinical Groups (FinnGen phases 1&2) | **Women's Health and Reproduction Group** |
| Ilkka Kalliala | Hospital District of Helsinki and Uusimaa, Helsinki, Finland | Clinical Groups (FinnGen phases 1&2) | **Women's Health and Reproduction Group** |
| Lauri Aaltonen | Hospital District of Helsinki and Uusimaa, Helsinki, Finland | Clinical Groups (FinnGen phases 1&2) | **Women's Health and Reproduction Group** |
| Varpu Jokimaa | Wellbeing Services County of Southwest Finland, Turku, Finland | Clinical Groups (FinnGen phases 1&2) | **Women's Health and Reproduction Group** |
| Johannes Kettunen | Northern Finland Biobank Borealis / University of Oulu / Wellbeing services county of North Ostrobothnia, Oulu, Finland | Clinical Groups (FinnGen phases 1&2) | **Women's Health and Reproduction Group** |

| | | | |
|---|---|---|---|
| Marja Vääräsmäki | Wellbeing services county of North Ostrobothnia, Oulu, Finland | Clinical Groups (FinnGen phases 1&2) | **Women's Health and Reproduction Group** |
| Outi Uimari | Wellbeing services county of North Ostrobothnia, Oulu, Finland | Clinical Groups (FinnGen phases 1&2) | **Women's Health and Reproduction Group** |
| Laure Morin-Papunen | Wellbeing services county of North Ostrobothnia, Oulu, Finland | Clinical Groups (FinnGen phases 1&2) | **Women's Health and Reproduction Group** |
| Maarit Niinimäki | Wellbeing services county of North Ostrobothnia, Oulu, Finland | Clinical Groups (FinnGen phases 1&2) | **Women's Health and Reproduction Group** |
| Terhi Piltonen | Wellbeing services county of North Ostrobothnia, Oulu, Finland | Clinical Groups (FinnGen phases 1&2) | **Women's Health and Reproduction Group** |
| Katja Kivinen | Institute for Molecular Medicine Finland (FIMM), HiLIFE, University of Helsinki, Helsinki, Finland | Clinical Groups (FinnGen phases 1&2) | **Women's Health and Reproduction Group** |
| Elisabeth Widen | Institute for Molecular Medicine Finland (FIMM), HiLIFE, University of Helsinki, Helsinki, Finland | Clinical Groups (FinnGen phases 1&2) | **Women's Health and Reproduction Group** |
| Taru Tukiainen | Institute for Molecular Medicine Finland (FIMM), HiLIFE, University of Helsinki, Helsinki, Finland | Clinical Groups (FinnGen phases 1&2) | **Women's Health and Reproduction Group** |
| Mary Pat Reeve | Institute for Molecular Medicine Finland (FIMM), HiLIFE, University of Helsinki, Helsinki, Finland | Clinical Groups (FinnGen phases 1&2) | **Women's Health and Reproduction Group** |
| Mark Daly | Institute for Molecular Medicine Finland (FIMM), HiLIFE, University of Helsinki, Helsinki, Finland; Broad Institute of MIT and Harvard; Massachusetts General Hospital, Boston, MA, United States | Clinical Groups (FinnGen phases 1&2) | **Women's Health and Reproduction Group** |
| Niko Välimäki | University of Helsinki, Helsinki, Finland | Clinical Groups (FinnGen phases 1&2) | **Women's Health and Reproduction Group** |
| Eija Laakkonen | University of Jyväskylä, Jyväskylä, Finland | Clinical Groups (FinnGen phases 1&2) | **Women's Health and Reproduction Group** |
| Jaakko Tyrmi | University of Oulu, Oulu, Finland / University of Tampere, Tampere, Finland | Clinical Groups (FinnGen phases 1&2) | **Women's Health and Reproduction Group** |
| Heidi Silven | University of Oulu, Oulu, Finland | Clinical Groups (FinnGen phases 1&2) | **Women's Health and Reproduction Group** |
| Eeva Sliz | University of Oulu, Oulu, Finland | Clinical Groups (FinnGen phases 1&2) | **Women's Health and Reproduction Group** |
| Riikka Arffman | University of Oulu, Oulu, Finland | Clinical Groups (FinnGen phases 1&2) | **Women's Health and Reproduction Group** |
| Susanna Savukoski | University of Oulu, Oulu, Finland | Clinical Groups (FinnGen phases 1&2) | **Women's Health and Reproduction Group** |
| Triin Laisk | Estonian biobank, Tartu, Estonia | Clinical Groups (FinnGen phases 1&2) | **Women's Health and Reproduction Group** |
| Natalia Pujol | Estonian biobank, Tartu, Estonia | Clinical Groups (FinnGen phases 1&2) | **Women's Health and Reproduction Group** |
| Mengzhen Liu | AbbVie, Chicago, IL, United States | Clinical Groups (FinnGen phases 1&2) | **Women's Health and Reproduction Group** |
| Bridget Riley-Gillis | AbbVie, Chicago, IL, United States | Clinical Groups (FinnGen phases 1&2) | **Women's Health and Reproduction Group** |
| Rion Pendergrass | Genentech, San Francisco, CA, United States | Clinical Groups (FinnGen phases 1&2) | **Women's Health and Reproduction Group** |
| Janet Kumar | GlaxoSmithKline, Collegeville, PA, United States | Clinical Groups (FinnGen phases 1&2) | **Women's Health and Reproduction Group** |
| Iiris Hovatta | University of Helsinki, Finland | Clinical Groups (FinnGen phases 1&2) | **Depression group** |

| | | | |
|---|---|---|---|
| Erkki Isometsä | Hospital District of Helsinki and Uusimaa, Helsinki, Finland | **Clinical Groups (FinnGen phases 1&2)** | **Depression group** |
| Hanna Ollila | Institute for Molecular Medicine Finland (FIMM), HiLIFE, University of Helsinki, Helsinki, Finland | **Clinical Groups (FinnGen phases 1&2)** | **Depression group** |
| Jaana Suvisaari | Finnish Institute for Health and Welfare (THL), Helsinki, Finland | **Clinical Groups (FinnGen phases 1&2)** | **Depression group** |
| Antti Mäkitie | University of Helsinki and Helsinki University Hospital, Helsinki, Finland | **Clinical Groups (FinnGen phases 1&2)** | **ENT (ear, nose and throath) Group** |
| Argyro Bizaki-Vallaskangas | Wellbeing Services County of Pirkanmaa, Tampere, Finland | **Clinical Groups (FinnGen phases 1&2)** | **ENT (ear, nose and throath) Group** |
| Sanna Toppila-Salmi | University of Eastern Finland and Kuopio University Hospital, Kuopio, Finland; Helsinki University Hospital and University of Helsinki, Finland | **Clinical Groups (FinnGen phases 1&2)** | **ENT (ear, nose and throath) Group** |
| Tytti Willberg | Wellbeing Services County of Southwest Finland, Turku, Finland | **Clinical Groups (FinnGen phases 1&2)** | **ENT (ear, nose and throath) Group** |
| Elmo Saarentaus | Institute for Molecular Medicine Finland (FIMM), HiLIFE, University of Helsinki, Helsinki, Finland | **Clinical Groups (FinnGen phases 1&2)** | **ENT (ear, nose and throath) Group** |
| Antti Aarnisalo | Hospital District of Helsinki and Uusimaa, Helsinki, Finland | **Clinical Groups (FinnGen phases 1&2)** | **ENT (ear, nose and throath) Group** |
| Eveliina Salminen | Hospital District of Helsinki and Uusimaa, Helsinki, Finland | **Clinical Groups (FinnGen phases 1&2)** | **ENT (ear, nose and throath) Group** |
| Elisa Rahikkala | Northern Ostrobothnia Hospital District, Oulu, Finland | **Clinical Groups (FinnGen phases 1&2)** | **ENT (ear, nose and throath) Group** |
| Johannes Kettunen | Northern Finland Biobank Borealis / University of Oulu / Wellbeing services county of North Ostrobothnia, Oulu, Finland | **Clinical Groups (FinnGen phases 1&2)** | **ENT (ear, nose and throath) Group** |
| Kristiina Aittomäki | Helsinki University Central Hospital, Helsinki, Finland | **Clinical Groups (FinnGen phases 1&2)** | **POI (premature ovarian failure) Group** |
| Fredrik Åberg | Helsinki University Hospital and University of Helsinki, Helsinki, Finland | **Clinical Groups (FinnGen phases 1&2)** | **LiverScore Group** |
| Joel Rämö | Institute for Molecular Medicine Finland (FIMM), HiLIFE, University of Helsinki, Helsinki, Finland; Broad Institute, Cambridge, MA, United States | **Clinical Task Forces (FinnGen phase 3)** | **Eye diseases Task Force** |
| Mark Daly | Institute for Molecular Medicine Finland (FIMM), HiLIFE, University of Helsinki, Helsinki, Finland; Broad Institute of MIT and Harvard; Massachusetts General Hospital, Boston, MA, United States | **Clinical Task Forces (FinnGen phase 3)** | **Eye diseases Task Force** |
| Mary Pat Reeve | Institute for Molecular Medicine Finland (FIMM), HiLIFE, University of Helsinki, Helsinki, Finland; Broad Institute, Cambridge, MA, United States | **Clinical Task Forces (FinnGen phase 3)** | **Eye diseases Task Force** |
| Muhammad Adnan Khan | Institute for Molecular Medicine Finland (FIMM), HiLIFE, University of Helsinki, Helsinki, Finland | **Clinical Task Forces (FinnGen phase 3)** | **Eye diseases Task Force** |
| Johanna Mäkelä | Finnish Biobank Cooperative - FINBB | **Clinical Task Forces (FinnGen phase 3)** | **Eye diseases Task Force** |
| Ilkka Immonen | Hospital District of Helsinki and Uusimaa, Helsinki, Finland | **Clinical Task Forces (FinnGen phase 3)** | **Eye diseases Task Force** |
| Kai Kaarniranta | Wellbeing services county of North Savo, Kuopio, Finland; University of Lodz, Lodz, Poland | **Clinical Task Forces (FinnGen phase 3)** | **Eye diseases Task Force** |
| Joni A Turunen | Helsinki University Hospital and University of Helsinki, Helsinki, Finland; Folkhälsan Research Center, Helsinki, Finland | **Clinical Task Forces (FinnGen phase 3)** | **Eye diseases Task Force** |
| Anneke Den Hollander | AbbVie, Chicago, IL, United States | **Clinical Task Forces (FinnGen phase 3)** | **Eye diseases Task Force** |
| Bridget Riley-Gillis | AbbVie, Chicago, IL, United States | **Clinical Task Forces (FinnGen phase 3)** | **Eye diseases Task Force** |

| | | | |
|---|---|---|---|
| Mengzhen Liu | AbbVie, Chicago, IL, United States | **Clinical Task Forces (FinnGen phase 3)** | **Eye diseases Task Force** |
| Nizar Smaoui | AbbVie, Chicago, IL, United States | **Clinical Task Forces (FinnGen phase 3)** | **Eye diseases Task Force** |
| Fabio Baschiera | Bayer AG, Leverkusen, Germany | **Clinical Task Forces (FinnGen phase 3)** | **Eye diseases Task Force** |
| Hans van Leeuwen | Bayer AG, Leverkusen, Germany | **Clinical Task Forces (FinnGen phase 3)** | **Eye diseases Task Force** |
| Elke Markert | Boehringer Ingelheim, Ingelheim am Rhein, Germany | **Clinical Task Forces (FinnGen phase 3)** | **Eye diseases Task Force** |
| Brian Yaspan | Genentech, San Francisco, CA, United States | **Clinical Task Forces (FinnGen phase 3)** | **Eye diseases Task Force** |
| Charli Harlow | GlaxoSmithKline, Collegeville, PA, United States | **Clinical Task Forces (FinnGen phase 3)** | **Eye diseases Task Force** |
| Lea Sarow-Blat | GlaxoSmithKline, Collegeville, PA, United States | **Clinical Task Forces (FinnGen phase 3)** | **Eye diseases Task Force** |
| Dermont Reilly | Johnson & Johnson Innovative Medicine, Spring House, PA, United States | **Clinical Task Forces (FinnGen phase 3)** | **Eye diseases Task Force** |
| P. Dunnmon | Johnson & Johnson Innovative Medicine, Spring House, PA, United States | **Clinical Task Forces (FinnGen phase 3)** | **Eye diseases Task Force** |
| Sara Gale | Johnson & Johnson Innovative Medicine, Spring House, PA, United States | **Clinical Task Forces (FinnGen phase 3)** | **Eye diseases Task Force** |
| Fabiana Farias | Merck, Kenilworth, NJ, United States | **Clinical Task Forces (FinnGen phase 3)** | **Eye diseases Task Force** |
| Jorge Del-aguila | Merck, Kenilworth, NJ, United States | **Clinical Task Forces (FinnGen phase 3)** | **Eye diseases Task Force** |
| Catherine O'Riordan | Translational Sciences, Sanofi R&D, Framingham, MA, USA | **Clinical Task Forces (FinnGen phase 3)** | **Eye diseases Task Force** |
| Samuel Lessard | Translational Sciences, Sanofi R&D, Framingham, MA, USA | **Clinical Task Forces (FinnGen phase 3)** | **Eye diseases Task Force** |
| Suzanne Jacobs | Translational Sciences, Sanofi R&D, Framingham, MA, USA | **Clinical Task Forces (FinnGen phase 3)** | **Eye diseases Task Force** |
| Satu Koskela | Finnish Red Cross Blood Service / Finnish Hematology Registry and Clinical Biobank, Helsinki, Finland | **Clinical Task Forces (FinnGen phase 3)** | **Immune mediated diseases Task Force** |
| Anne Kerola | Institute for Molecular Medicine Finland (FIMM), HiLIFE, University of Helsinki, Helsinki, Finland | **Clinical Task Forces (FinnGen phase 3)** | **Immune mediated diseases Task Force** |
| Elisa Lahtela | Institute for Molecular Medicine Finland (FIMM), HiLIFE, University of Helsinki, Helsinki, Finland | **Clinical Task Forces (FinnGen phase 3)** | **Immune mediated diseases Task Force** |
| Helen Cooper | Institute for Molecular Medicine Finland (FIMM), HiLIFE, University of Helsinki, Helsinki, Finland | **Clinical Task Forces (FinnGen phase 3)** | **Immune mediated diseases Task Force** |
| Johanna Paltta | Institute for Molecular Medicine Finland (FIMM), HiLIFE, University of Helsinki, Helsinki, Finland; University of Turku, Turku, Finland | **Clinical Task Forces (FinnGen phase 3)** | **Immune mediated diseases Task Force** |
| Jukka Koskela | Institute for Molecular Medicine Finland (FIMM), HiLIFE, University of Helsinki, Helsinki, Finland | **Clinical Task Forces (FinnGen phase 3)** | **Immune mediated diseases Task Force** |
| Mark Daly | Institute for Molecular Medicine Finland (FIMM), HiLIFE, University of Helsinki, Helsinki, Finland; Broad Institute of MIT and Harvard; Massachusetts General Hospital, Boston, MA, United States | **Clinical Task Forces (FinnGen phase 3)** | **Immune mediated diseases Task Force** |
| Mary Pat Reeve | Institute for Molecular Medicine Finland (FIMM), HiLIFE, University of Helsinki, Helsinki, Finland; Broad Institute, Cambridge, MA, United States | **Clinical Task Forces (FinnGen phase 3)** | **Immune mediated diseases Task Force** |

| | | | |
|---|---|---|---|
| Vincent Llorens | Institute for Molecular Medicine Finland (FIMM), HiLIFE, University of Helsinki, Helsinki, Finland | **Clinical Task Forces (FinnGen phase 3)** | **Immune mediated diseases Task Force** |
| Martti Färkkilä | Hospital District of Helsinki and Uusimaa, Helsinki, Finland | **Clinical Task Forces (FinnGen phase 3)** | **Immune mediated diseases Task Force** |
| Johannes Kettunen | Northern Finland Biobank Borealis / University of Oulu / Wellbeing services county of North Ostrobothnia, Oulu, Finland | **Clinical Task Forces (FinnGen phase 3)** | **Immune mediated diseases Task Force** |
| Kaisa Tasanen-Maatta | Wellbeing services county of North Ostrobothnia, Oulu, Finland | **Clinical Task Forces (FinnGen phase 3)** | **Immune mediated diseases Task Force** |
| Laura Huilaja | Wellbeing services county of North Ostrobothnia, Oulu, Finland | **Clinical Task Forces (FinnGen phase 3)** | **Immune mediated diseases Task Force** |
| Minna Ruddock | Arctic biobank / University of Oulu, Oulu, Finland | **Clinical Task Forces (FinnGen phase 3)** | **Immune mediated diseases Task Force** |
| Aki Havulinna | Finnish Institute for Health and Welfare (THL), Helsinki, Finland | **Clinical Task Forces (FinnGen phase 3)** | **Immune mediated diseases Task Force** |
| Antti Palomäki | University of Turku, Turku, Finland | **Clinical Task Forces (FinnGen phase 3)** | **Immune mediated diseases Task Force** |
| Laura Kuusalo | University of Turku, Turku, Finland | **Clinical Task Forces (FinnGen phase 3)** | **Immune mediated diseases Task Force** |
| Laura Pirilä | University of Turku, Turku, Finland | **Clinical Task Forces (FinnGen phase 3)** | **Immune mediated diseases Task Force** |
| Fedik Rahimov | AbbVie, Chicago, IL, United States | **Clinical Task Forces (FinnGen phase 3)** | **Immune mediated diseases Task Force** |
| Jan Freudenberg | AbbVie, Chicago, IL, United States | **Clinical Task Forces (FinnGen phase 3)** | **Immune mediated diseases Task Force** |
| Nizar Smaoui | AbbVie, Chicago, IL, United States | **Clinical Task Forces (FinnGen phase 3)** | **Immune mediated diseases Task Force** |
| Bram Prins | Astra Zeneca, Cambridge, United Kingdom | **Clinical Task Forces (FinnGen phase 3)** | **Immune mediated diseases Task Force** |
| Coralie Viollet | Astra Zeneca, Cambridge, United Kingdom | **Clinical Task Forces (FinnGen phase 3)** | **Immune mediated diseases Task Force** |
| Eleanor Wheeler | Astra Zeneca, Cambridge, United Kingdom | **Clinical Task Forces (FinnGen phase 3)** | **Immune mediated diseases Task Force** |
| Kousik Kundu | Astra Zeneca, Cambridge, United Kingdom | **Clinical Task Forces (FinnGen phase 3)** | **Immune mediated diseases Task Force** |
| Santosh Atanur | Astra Zeneca, Cambridge, United Kingdom | **Clinical Task Forces (FinnGen phase 3)** | **Immune mediated diseases Task Force** |
| Hans van Leeuwen | Bayer AG, Leverkusen, Germany | **Clinical Task Forces (FinnGen phase 3)** | **Immune mediated diseases Task Force** |
| Himanshu Manchanda | Bayer AG, Leverkusen, Germany | **Clinical Task Forces (FinnGen phase 3)** | **Immune mediated diseases Task Force** |
| Karl Heilbron | Bayer AG, Leverkusen, Germany | **Clinical Task Forces (FinnGen phase 3)** | **Immune mediated diseases Task Force** |
| Martin Rao | Bayer AG, Leverkusen, Germany | **Clinical Task Forces (FinnGen phase 3)** | **Immune mediated diseases Task Force** |
| Nicole Schmidt | Bayer AG, Leverkusen, Germany | **Clinical Task Forces (FinnGen phase 3)** | **Immune mediated diseases Task Force** |
| Samu Kurki | Bayer AG, Leverkusen, Germany | **Clinical Task Forces (FinnGen phase 3)** | **Immune mediated diseases Task Force** |

| | | | |
|---|---|---|---|
| Ellen Tsai | Biogen, Cambridge, MA, United States | Clinical Task Forces (FinnGen phase 3) | **Immune mediated diseases Task Force** |
| Ketian Yu | Biogen, Cambridge, MA, United States | Clinical Task Forces (FinnGen phase 3) | **Immune mediated diseases Task Force** |
| Stephanie Loomis | Biogen, Cambridge, MA, United States | Clinical Task Forces (FinnGen phase 3) | **Immune mediated diseases Task Force** |
| Benjamin Sun | Bristol Myers Squibb, New York, NY, United States | Clinical Task Forces (FinnGen phase 3) | **Immune mediated diseases Task Force** |
| Cara Carty | Bristol Myers Squibb, New York, NY, United States | Clinical Task Forces (FinnGen phase 3) | **Immune mediated diseases Task Force** |
| Emily Holzinger | Bristol Myers Squibb, New York, NY, United States | Clinical Task Forces (FinnGen phase 3) | **Immune mediated diseases Task Force** |
| Michael Turchin | Bristol Myers Squibb, New York, NY, United States | Clinical Task Forces (FinnGen phase 3) | **Immune mediated diseases Task Force** |
| Neelakshi Jog | Bristol Myers Squibb, New York, NY, United States | Clinical Task Forces (FinnGen phase 3) | **Immune mediated diseases Task Force** |
| Frank Li | Boehringer Ingelheim, Ingelheim am Rhein, Germany | Clinical Task Forces (FinnGen phase 3) | **Immune mediated diseases Task Force** |
| Zhihao Ding | Boehringer Ingelheim, Ingelheim am Rhein, Germany | Clinical Task Forces (FinnGen phase 3) | **Immune mediated diseases Task Force** |
| Cameron Adams | Genentech, San Francisco, CA, United States | Clinical Task Forces (FinnGen phase 3) | **Immune mediated diseases Task Force** |
| Mark McCarthy | Genentech, San Francisco, CA, United States | Clinical Task Forces (FinnGen phase 3) | **Immune mediated diseases Task Force** |
| Michael Rothenberg | Genentech, San Francisco, CA, United States | Clinical Task Forces (FinnGen phase 3) | **Immune mediated diseases Task Force** |
| Rion Pendergrass | Genentech, San Francisco, CA, United States | Clinical Task Forces (FinnGen phase 3) | **Immune mediated diseases Task Force** |
| Diana L.Cousminer | GlaxoSmithKline, Collegeville, PA, United States | Clinical Task Forces (FinnGen phase 3) | **Immune mediated diseases Task Force** |
| Jagtar Nijjar | GlaxoSmithKline, Collegeville, PA, United States | Clinical Task Forces (FinnGen phase 3) | **Immune mediated diseases Task Force** |
| Jessica Chao | GlaxoSmithKline, Collegeville, PA, United States | Clinical Task Forces (FinnGen phase 3) | **Immune mediated diseases Task Force** |
| Joanna C.Betts | GlaxoSmithKline, Collegeville, PA, United States | Clinical Task Forces (FinnGen phase 3) | **Immune mediated diseases Task Force** |
| Jonathan M.Davitte | GlaxoSmithKline, Collegeville, PA, United States | Clinical Task Forces (FinnGen phase 3) | **Immune mediated diseases Task Force** |
| Linda McGarthy | GlaxoSmithKline, Collegeville, PA, United States | Clinical Task Forces (FinnGen phase 3) | **Immune mediated diseases Task Force** |
| Michal Magid | GlaxoSmithKline, Collegeville, PA, United States | Clinical Task Forces (FinnGen phase 3) | **Immune mediated diseases Task Force** |
| Shashank Jariwala | GlaxoSmithKline, Collegeville, PA, United States | Clinical Task Forces (FinnGen phase 3) | **Immune mediated diseases Task Force** |
| Dawn Waterworth | Johnson & Johnson Innovative Medicine, Spring House, PA, United States | Clinical Task Forces (FinnGen phase 3) | **Immune mediated diseases Task Force** |
| Amy Hart | Johnson & Johnson Innovative Medicine, Spring House, PA, United States | Clinical Task Forces (FinnGen phase 3) | **Immune mediated diseases Task Force** |

| | | | |
|---|---|---|---|
| Brice Keyes | Johnson & Johnson Innovative Medicine, Spring House, PA, United States | **Clinical Task Forces (FinnGen phase 3)** | **Immune mediated diseases Task Force** |
| John Kwon | Johnson & Johnson Innovative Medicine, Spring House, PA, United States | **Clinical Task Forces (FinnGen phase 3)** | **Immune mediated diseases Task Force** |
| Jonathan Sherlock | Johnson & Johnson Innovative Medicine, Spring House, PA, United States | **Clinical Task Forces (FinnGen phase 3)** | **Immune mediated diseases Task Force** |
| Matt Loza | Johnson & Johnson Innovative Medicine, Spring House, PA, United States | **Clinical Task Forces (FinnGen phase 3)** | **Immune mediated diseases Task Force** |
| Elisabeth Vollmann | Merck, Kenilworth, NJ, United States | **Clinical Task Forces (FinnGen phase 3)** | **Immune mediated diseases Task Force** |
| Jozsef Karman | Merck, Kenilworth, NJ, United States | **Clinical Task Forces (FinnGen phase 3)** | **Immune mediated diseases Task Force** |
| Julie Fiore | Merck, Kenilworth, NJ, United States | **Clinical Task Forces (FinnGen phase 3)** | **Immune mediated diseases Task Force** |
| Rajesh Kamath | Merck, Kenilworth, NJ, United States | **Clinical Task Forces (FinnGen phase 3)** | **Immune mediated diseases Task Force** |
| Enrico Ferrero | Novartis Institutes for BioMedical Research, Cambridge, MA, United States | **Clinical Task Forces (FinnGen phase 3)** | **Immune mediated diseases Task Force** |
| Jonas Zierer | Novartis Institutes for BioMedical Research, Cambridge, MA, United States | **Clinical Task Forces (FinnGen phase 3)** | **Immune mediated diseases Task Force** |
| Nikos Patsopoulos | Novartis Institutes for BioMedical Research, Cambridge, MA, United States | **Clinical Task Forces (FinnGen phase 3)** | **Immune mediated diseases Task Force** |
| Erin Macdonald-Dunlop | Pfizer, New York, NY, United States | **Clinical Task Forces (FinnGen phase 3)** | **Immune mediated diseases Task Force** |
| Jessica Chung | Pfizer, New York, NY, United States | **Clinical Task Forces (FinnGen phase 3)** | **Immune mediated diseases Task Force** |
| Michael McLean | Pfizer, New York, NY, United States | **Clinical Task Forces (FinnGen phase 3)** | **Immune mediated diseases Task Force** |
| Hamid Mattoo | Translational Sciences, Sanofi R&D, Framingham, MA, USA | **Clinical Task Forces (FinnGen phase 3)** | **Immune mediated diseases Task Force** |
| Aarno Palotie | Institute for Molecular Medicine Finland (FIMM), HiLIFE, University of Helsinki, Helsinki, Finland; Broad Institute of MIT and Harvard; Massachusetts General Hospital, Boston, MA, United States | **Clinical Task Forces (FinnGen phase 3)** | **Kidney diseases Task Force** |
| Elisa Lahtela | Institute for Molecular Medicine Finland (FIMM), HiLIFE, University of Helsinki, Helsinki, Finland | **Clinical Task Forces (FinnGen phase 3)** | **Kidney diseases Task Force** |
| Helen Cooper | Institute for Molecular Medicine Finland (FIMM), HiLIFE, University of Helsinki, Helsinki, Finland | **Clinical Task Forces (FinnGen phase 3)** | **Kidney diseases Task Force** |
| Jukka Koskela | Institute for Molecular Medicine Finland (FIMM), HiLIFE, University of Helsinki, Helsinki, Finland | **Clinical Task Forces (FinnGen phase 3)** | **Kidney diseases Task Force** |
| Mark Daly | Institute for Molecular Medicine Finland (FIMM), HiLIFE, University of Helsinki, Helsinki, Finland; Broad Institute of MIT and Harvard; Massachusetts General Hospital, Boston, MA, United States | **Clinical Task Forces (FinnGen phase 3)** | **Kidney diseases Task Force** |
| Mary Pat Reeve | Institute for Molecular Medicine Finland (FIMM), HiLIFE, University of Helsinki, Helsinki, Finland; Broad Institute, Cambridge, MA, United States | **Clinical Task Forces (FinnGen phase 3)** | **Kidney diseases Task Force** |
| Raymond Walters | Institute for Molecular Medicine Finland (FIMM), HiLIFE, University of Helsinki, Helsinki, Finland; Broad Institute, Cambridge, MA, United States | **Clinical Task Forces (FinnGen phase 3)** | **Kidney diseases Task Force** |
| Rodos Rodosthenous | Institute for Molecular Medicine Finland (FIMM), HiLIFE, University of Helsinki, Helsinki, Finland | **Clinical Task Forces (FinnGen phase 3)** | **Kidney diseases Task Force** |
| Jouni Lauronen | Finnish Red Cross Blood Service / Finnish Hematology Registry and Clinical Biobank, Helsinki, Finland | **Clinical Task Forces (FinnGen phase 3)** | **Kidney diseases Task Force** |

| | | | |
|---|---|---|---|
| Adrian Banerji | Institute for Molecular Medicine Finland (FIMM), HiLIFE, University of Helsinki, Helsinki, Finland; Broad Institute & Harvard Medical School, Cambridge, United States | Clinical Task Forces (FinnGen phase 3) | **Kidney diseases Task Force** |
| Matthew Sampson | Broad Institute, Cambridge, MA, United States; Harvard Medical School, Cambridge, United States | Clinical Task Forces (FinnGen phase 3) | **Kidney diseases Task Force** |
| Michelle McNulty | Institute for Molecular Medicine Finland (FIMM), HiLIFE, University of Helsinki, Helsinki, Finland; Broad Institute & Harvard Medical School, Cambridge, United States | Clinical Task Forces (FinnGen phase 3) | **Kidney diseases Task Force** |
| Daniel Gordin | Helsinki University Hospital and University of Helsinki, Helsinki, Finland | Clinical Task Forces (FinnGen phase 3) | **Kidney diseases Task Force** |
| Patrik Finne | Helsinki University Hospital and University of Helsinki, Helsinki, Finland | Clinical Task Forces (FinnGen phase 3) | **Kidney diseases Task Force** |
| Mika Kähönen | Finnish Clinical Biobank Tampere / University of Tampere / Wellbeing Services County of Pirkanmaa, Tampere, Finland | Clinical Task Forces (FinnGen phase 3) | **Kidney diseases Task Force** |
| Tapio Hellman | University of Turku, Turku, Finland | Clinical Task Forces (FinnGen phase 3) | **Kidney diseases Task Force** |
| Teemu Niiranen | University of Turku, Turku, Finland; Finnish Institute for Health and Welfare (THL), Helsinki, Finland | Clinical Task Forces (FinnGen phase 3) | **Kidney diseases Task Force** |
| Dirk Paul | Astra Zeneca, Cambridge, United Kingdom | Clinical Task Forces (FinnGen phase 3) | **Kidney diseases Task Force** |
| Ioanna Tachmazidou | Astra Zeneca, Cambridge, United Kingdom | Clinical Task Forces (FinnGen phase 3) | **Kidney diseases Task Force** |
| Hans van Leeuwen | Bayer AG, Leverkusen, Germany | Clinical Task Forces (FinnGen phase 3) | **Kidney diseases Task Force** |
| Johanna Mielke | Bayer AG, Leverkusen, Germany | Clinical Task Forces (FinnGen phase 3) | **Kidney diseases Task Force** |
| Juho Immonen | Bayer AG, Leverkusen, Germany | Clinical Task Forces (FinnGen phase 3) | **Kidney diseases Task Force** |
| Thomas Battram | Bayer AG, Leverkusen, Germany | Clinical Task Forces (FinnGen phase 3) | **Kidney diseases Task Force** |
| Tobias Hogrebe | Bayer AG, Leverkusen, Germany | Clinical Task Forces (FinnGen phase 3) | **Kidney diseases Task Force** |
| Ketian Yu | Biogen, Cambridge, MA, United States | Clinical Task Forces (FinnGen phase 3) | **Kidney diseases Task Force** |
| Benjamin Sun | Bristol Myers Squibb, New York, NY, United States | Clinical Task Forces (FinnGen phase 3) | **Kidney diseases Task Force** |
| Janie Shelton | Bristol Myers Squibb, New York, NY, United States | Clinical Task Forces (FinnGen phase 3) | **Kidney diseases Task Force** |
| Yao Hu | Boehringer Ingelheim, Ingelheim am Rhein, Germany | Clinical Task Forces (FinnGen phase 3) | **Kidney diseases Task Force** |
| Zhihao Ding | Boehringer Ingelheim, Ingelheim am Rhein, Germany | Clinical Task Forces (FinnGen phase 3) | **Kidney diseases Task Force** |
| Rion Pendergrass | Genentech, San Francisco, CA, United States | Clinical Task Forces (FinnGen phase 3) | **Kidney diseases Task Force** |
| Sergio Dellepiane | Genentech, San Francisco, CA, United States | Clinical Task Forces (FinnGen phase 3) | **Kidney diseases Task Force** |
| Audrey Chu | GlaxoSmithKline, Collegeville, PA, United States | Clinical Task Forces (FinnGen phase 3) | **Kidney diseases Task Force** |
| Chris Floyd | GlaxoSmithKline, Collegeville, PA, United States | Clinical Task Forces (FinnGen phase 3) | **Kidney diseases Task Force** |

| | | | |
|---|---|---|---|
| Dan Swerdlow | GlaxoSmithKline, Collegeville, PA, United States | **Clinical Task Forces (FinnGen phase 3)** | **Kidney diseases Task Force** |
| Erding Hu | GlaxoSmithKline, Collegeville, PA, United States | **Clinical Task Forces (FinnGen phase 3)** | **Kidney diseases Task Force** |
| Jonathan Davitte | GlaxoSmithKline, Collegeville, PA, United States | **Clinical Task Forces (FinnGen phase 3)** | **Kidney diseases Task Force** |
| Prerak Desai | GlaxoSmithKline, Collegeville, PA, United States | **Clinical Task Forces (FinnGen phase 3)** | **Kidney diseases Task Force** |
| Stephen Haddad | GlaxoSmithKline, Collegeville, PA, United States | **Clinical Task Forces (FinnGen phase 3)** | **Kidney diseases Task Force** |
| Dermot Reilly | Johnson & Johnson Innovative Medicine, Spring House, PA, United States | **Clinical Task Forces (FinnGen phase 3)** | **Kidney diseases Task Force** |
| P. Dunnmon | Johnson & Johnson Innovative Medicine, Spring House, PA, United States | **Clinical Task Forces (FinnGen phase 3)** | **Kidney diseases Task Force** |
| Karol Estrada | Maze Therapeutics, San Francisco, CA, United States | **Clinical Task Forces (FinnGen phase 3)** | **Kidney diseases Task Force** |
| Rob Graham | Maze Therapeutics, San Francisco, CA, United States | **Clinical Task Forces (FinnGen phase 3)** | **Kidney diseases Task Force** |
| Sahar Mozzafari | Maze Therapeutics, San Francisco, CA, United States | **Clinical Task Forces (FinnGen phase 3)** | **Kidney diseases Task Force** |
| Nancy Finkel | Novartis Institutes for BioMedical Research, Cambridge, MA, United States | **Clinical Task Forces (FinnGen phase 3)** | **Kidney diseases Task Force** |
| Sabina Pfister | Novartis Institutes for BioMedical Research, Cambridge, MA, United States | **Clinical Task Forces (FinnGen phase 3)** | **Kidney diseases Task Force** |
| Shola Richards | Novartis Institutes for BioMedical Research, Cambridge, MA, United States | **Clinical Task Forces (FinnGen phase 3)** | **Kidney diseases Task Force** |
| Joshua Chiou | Pfizer, New York, NY, United States | **Clinical Task Forces (FinnGen phase 3)** | **Kidney diseases Task Force** |
| Ying Wu | Pfizer, New York, NY, United States | **Clinical Task Forces (FinnGen phase 3)** | **Kidney diseases Task Force** |
| Katherine Klinger | Translational Sciences, Sanofi R&D, Framingham, MA, USA | **Clinical Task Forces (FinnGen phase 3)** | **Kidney diseases Task Force** |
| Matti Vuori | University of Turku, Turku, Finland | **Clinical Task Forces (FinnGen phase 3)** | **Metabolic diseases Task Force** |
| Teemu Niiranen | University of Turku, Turku, Finland; Finnish Institute for Health and Welfare (THL), Helsinki, Finland | **Clinical Task Forces (FinnGen phase 3)** | **Metabolic diseases Task Force** |
| Bridget Riley-Gillis | AbbVie, Chicago, IL, United States | **Clinical Task Forces (FinnGen phase 3)** | **Metabolic diseases Task Force** |
| Nizar Smaoui | AbbVie, Chicago, IL, United States | **Clinical Task Forces (FinnGen phase 3)** | **Metabolic diseases Task Force** |
| Alix Berton | Bayer AG, Leverkusen, Germany | **Clinical Task Forces (FinnGen phase 3)** | **Metabolic diseases Task Force** |
| Hans van Leeuwen | Bayer AG, Leverkusen, Germany | **Clinical Task Forces (FinnGen phase 3)** | **Metabolic diseases Task Force** |
| Chen Li | Bristol Myers Squibb, New York, NY, United States | **Clinical Task Forces (FinnGen phase 3)** | **Metabolic diseases Task Force** |
| Emily Holzinger | Bristol Myers Squibb, New York, NY, United States | **Clinical Task Forces (FinnGen phase 3)** | **Metabolic diseases Task Force** |

| | | | |
|---|---|---|---|
| Anubha Mahajan | Genentech, San Francisco, CA, United States | Clinical Task Forces (FinnGen phase 3) | **Metabolic diseases Task Force** |
| Mark Mccarthy | Genentech, San Francisco, CA, United States | Clinical Task Forces (FinnGen phase 3) | **Metabolic diseases Task Force** |
| Christopher Deboever | Maze Therapeutics, San Francisco, CA, United States | Clinical Task Forces (FinnGen phase 3) | **Metabolic diseases Task Force** |
| Karol Estrada | Maze Therapeutics, San Francisco, CA, United States | Clinical Task Forces (FinnGen phase 3) | **Metabolic diseases Task Force** |
| Robert Graham | Maze Therapeutics, San Francisco, CA, United States | Clinical Task Forces (FinnGen phase 3) | **Metabolic diseases Task Force** |
| Hye In Kim | Pfizer, New York, NY, United States | Clinical Task Forces (FinnGen phase 3) | **Metabolic diseases Task Force** |
| Sivakumar Pitchumani | Pfizer, New York, NY, United States | Clinical Task Forces (FinnGen phase 3) | **Metabolic diseases Task Force** |
| Sumedha Jassal | Pfizer, New York, NY, United States | Clinical Task Forces (FinnGen phase 3) | **Metabolic diseases Task Force** |
| Åsa Hedman | Pfizer, New York, NY, United States | Clinical Task Forces (FinnGen phase 3) | **Metabolic diseases Task Force** |
| Aarno Palotie | Institute for Molecular Medicine Finland (FIMM), HiLIFE, University of Helsinki, Helsinki, Finland; Broad Institute of MIT and Harvard; Massachusetts General Hospital, Boston, MA, United States | Clinical Task Forces (FinnGen phase 3) | **Neurodegenerative diseases Task Force** |
| Austin Argentieri | Institute for Molecular Medicine Finland (FIMM), HiLIFE, University of Helsinki, Helsinki, Finland; Broad Institute, Cambridge, MA, United States | Clinical Task Forces (FinnGen phase 3) | **Neurodegenerative diseases Task Force** |
| Aoxing Liu | Institute for Molecular Medicine Finland (FIMM), HiLIFE, University of Helsinki, Helsinki, Finland | Clinical Task Forces (FinnGen phase 3) | **Neurodegenerative diseases Task Force** |
| Eero Vuoksimaa | Institute for Molecular Medicine Finland (FIMM), HiLIFE, University of Helsinki, Helsinki, Finland | Clinical Task Forces (FinnGen phase 3) | **Neurodegenerative diseases Task Force** |
| Elisa Lahtela | Institute for Molecular Medicine Finland (FIMM), HiLIFE, University of Helsinki, Helsinki, Finland | Clinical Task Forces (FinnGen phase 3) | **Neurodegenerative diseases Task Force** |
| Joni Lindbohm | Institute for Molecular Medicine Finland (FIMM), HiLIFE, University of Helsinki, Helsinki, Finland | Clinical Task Forces (FinnGen phase 3) | **Neurodegenerative diseases Task Force** |
| Mark Daly | Institute for Molecular Medicine Finland (FIMM), HiLIFE, University of Helsinki, Helsinki, Finland; Broad Institute of MIT and Harvard; Massachusetts General Hospital, Boston, MA, United States | Clinical Task Forces (FinnGen phase 3) | **Neurodegenerative diseases Task Force** |
| Mary Pat Reeve | Institute for Molecular Medicine Finland (FIMM), HiLIFE, University of Helsinki, Helsinki, Finland; Broad Institute, Cambridge, MA, United States | Clinical Task Forces (FinnGen phase 3) | **Neurodegenerative diseases Task Force** |
| Paavo Häppölä | Institute for Molecular Medicine Finland (FIMM), HiLIFE, University of Helsinki, Helsinki, Finland | Clinical Task Forces (FinnGen phase 3) | **Neurodegenerative diseases Task Force** |
| Zhiyu Yang | Institute for Molecular Medicine Finland (FIMM), HiLIFE, University of Helsinki, Helsinki, Finland | Clinical Task Forces (FinnGen phase 3) | **Neurodegenerative diseases Task Force** |
| Eino Solje | University of Eastern Finland, Kuopio, Finland | Clinical Task Forces (FinnGen phase 3) | **Neurodegenerative diseases Task Force** |
| Mikko Hiltunen | University of Eastern Finland, Kuopio, Finland | Clinical Task Forces (FinnGen phase 3) | **Neurodegenerative diseases Task Force** |
| Valtteri Julkunen | University of Eastern Finland and Kuopio University Hospital, Kuopio, Finland | Clinical Task Forces (FinnGen phase 3) | **Neurodegenerative diseases Task Force** |
| Ville Leinonen | University of Eastern Finland and Kuopio University Hospital, Kuopio, Finland | Clinical Task Forces (FinnGen phase 3) | **Neurodegenerative diseases Task Force** |
| Hanna Kujala | Biobank of Eastern Finland / University of Eastern Finland / Wellbeing services county of North Savo, Kuopio, Finland | Clinical Task Forces (FinnGen phase 3) | **Neurodegenerative diseases Task Force** |

| | | | |
|---|---|---|---|
| Aki Havulinna | Finnish Institute for Health and Welfare (THL), Helsinki, Finland | **Clinical Task Forces (FinnGen phase 3)** | **Neurodegenerative diseases Task Force** |
| Roosa Kallionpää | University of Turku, Turku, Finland | **Clinical Task Forces (FinnGen phase 3)** | **Neurodegenerative diseases Task Force** |
| Minttu Marttila | University of Helsinki, Helsinki, Finland | **Clinical Task Forces (FinnGen phase 3)** | **Neurodegenerative diseases Task Force** |
| Britney Milkovich | AbbVie, Chicago, IL, United States | **Clinical Task Forces (FinnGen phase 3)** | **Neurodegenerative diseases Task Force** |
| Jan Freudenberg | AbbVie, Chicago, IL, United States | **Clinical Task Forces (FinnGen phase 3)** | **Neurodegenerative diseases Task Force** |
| Andrew Lowe | Astra Zeneca, Cambridge, United Kingdom | **Clinical Task Forces (FinnGen phase 3)** | **Neurodegenerative diseases Task Force** |
| Ioanna Tachmazidou | Astra Zeneca, Cambridge, United Kingdom | **Clinical Task Forces (FinnGen phase 3)** | **Neurodegenerative diseases Task Force** |
| Thomas Spargo | Astra Zeneca, Cambridge, United Kingdom | **Clinical Task Forces (FinnGen phase 3)** | **Neurodegenerative diseases Task Force** |
| Kritika Singh | Bristol Myers Squibb, New York, NY, United States | **Clinical Task Forces (FinnGen phase 3)** | **Neurodegenerative diseases Task Force** |
| Peng Jiang | Bristol Myers Squibb, New York, NY, United States | **Clinical Task Forces (FinnGen phase 3)** | **Neurodegenerative diseases Task Force** |
| Stephanie Loomis | Bristol Myers Squibb, New York, NY, United States | **Clinical Task Forces (FinnGen phase 3)** | **Neurodegenerative diseases Task Force** |
| Anubha Mahajan | Genentech, San Francisco, CA, United States | **Clinical Task Forces (FinnGen phase 3)** | **Neurodegenerative diseases Task Force** |
| Rion Pendergrass | Genentech, San Francisco, CA, United States | **Clinical Task Forces (FinnGen phase 3)** | **Neurodegenerative diseases Task Force** |
| Damien Croteau-Chonka | GlaxoSmithKline, Collegeville, PA, United States | **Clinical Task Forces (FinnGen phase 3)** | **Neurodegenerative diseases Task Force** |
| John Eicher | GlaxoSmithKline, Collegeville, PA, United States | **Clinical Task Forces (FinnGen phase 3)** | **Neurodegenerative diseases Task Force** |
| Prerak Desai | GlaxoSmithKline, Collegeville, PA, United States | **Clinical Task Forces (FinnGen phase 3)** | **Neurodegenerative diseases Task Force** |
| Chris Whelan | Johnson & Johnson Innovative Medicine, Spring House, PA, United States | **Clinical Task Forces (FinnGen phase 3)** | **Neurodegenerative diseases Task Force** |
| Karen He | Johnson & Johnson Innovative Medicine, Spring House, PA, United States | **Clinical Task Forces (FinnGen phase 3)** | **Neurodegenerative diseases Task Force** |
| Qingqin Li | Johnson & Johnson Innovative Medicine, Spring House, PA, United States | **Clinical Task Forces (FinnGen phase 3)** | **Neurodegenerative diseases Task Force** |
| W Galpern | Johnson & Johnson Innovative Medicine, Spring House, PA, United States | **Clinical Task Forces (FinnGen phase 3)** | **Neurodegenerative diseases Task Force** |
| Yanfei Zhang | Johnson & Johnson Innovative Medicine, Spring House, PA, United States | **Clinical Task Forces (FinnGen phase 3)** | **Neurodegenerative diseases Task Force** |
| Andrei Popescu | Merck, Kenilworth, NJ, United States | **Clinical Task Forces (FinnGen phase 3)** | **Neurodegenerative diseases Task Force** |
| Delphine Fagegaltier | Merck, Kenilworth, NJ, United States | **Clinical Task Forces (FinnGen phase 3)** | **Neurodegenerative diseases Task Force** |
| Mari Niemi | Novartis Institutes for BioMedical Research, Cambridge, MA, United States | **Clinical Task Forces (FinnGen phase 3)** | **Neurodegenerative diseases Task Force** |

| | | | |
|---|---|---|---|
| Nikos Patsopoulos | Novartis Institutes for BioMedical Research, Cambridge, MA, United States | **Clinical Task Forces (FinnGen phase 3)** | **Neurodegenerative diseases Task Force** |
| Katherine Klinger | Translational Sciences, Sanofi R&D, Framingham, MA, USA | **Clinical Task Forces (FinnGen phase 3)** | **Neurodegenerative diseases Task Force** |
| Aarno Palotie | Institute for Molecular Medicine Finland (FIMM), HiLIFE, University of Helsinki, Helsinki, Finland; Broad Institute of MIT and Harvard; Massachusetts General Hospital, Boston, MA, United States | **Clinical Task Forces (FinnGen phase 3)** | **Pulmonology Task Force and fibrotic diseases interest group** |
| Elisa Lahtela | Institute for Molecular Medicine Finland (FIMM), HiLIFE, University of Helsinki, Helsinki, Finland | **Clinical Task Forces (FinnGen phase 3)** | **Pulmonology Task Force and fibrotic diseases interest group** |
| Jukka Koskela | Institute for Molecular Medicine Finland (FIMM), HiLIFE, University of Helsinki, Helsinki, Finland | **Clinical Task Forces (FinnGen phase 3)** | **Pulmonology Task Force and fibrotic diseases interest group** |
| Mark Daly | Institute for Molecular Medicine Finland (FIMM), HiLIFE, University of Helsinki, Helsinki, Finland; Broad Institute of MIT and Harvard; Massachusetts General Hospital, Boston, MA, United States | **Clinical Task Forces (FinnGen phase 3)** | **Pulmonology Task Force and fibrotic diseases interest group** |
| Sanni Ruotsalainen | Institute for Molecular Medicine Finland (FIMM), HiLIFE, University of Helsinki, Helsinki, Finland | **Clinical Task Forces (FinnGen phase 3)** | **Pulmonology Task Force and fibrotic diseases interest group** |
| Susanna Lemmelä | Institute for Molecular Medicine Finland (FIMM), HiLIFE, University of Helsinki, Helsinki, Finland | **Clinical Task Forces (FinnGen phase 3)** | **Pulmonology Task Force and fibrotic diseases interest group** |
| Tarja Laitinen | Institute for Molecular Medicine Finland (FIMM), HiLIFE, University of Helsinki, Helsinki, Finland | **Clinical Task Forces (FinnGen phase 3)** | **Pulmonology Task Force and fibrotic diseases interest group** |
| Salla Ranta | Hospital District of Helsinki and Uusimaa, Helsinki, Finland | **Clinical Task Forces (FinnGen phase 3)** | **Pulmonology Task Force and fibrotic diseases interest group** |
| Paavo Häppölä | Institute for Molecular Medicine Finland (FIMM), HiLIFE, University of Helsinki, Helsinki, Finland | **Clinical Task Forces (FinnGen phase 3)** | **Pulmonology Task Force and fibrotic diseases interest group** |
| Paula Kauppi | Hospital District of Helsinki and Uusimaa, Helsinki, Finland | **Clinical Task Forces (FinnGen phase 3)** | **Pulmonology Task Force and fibrotic diseases interest group** |
| Tiinamaija Tuomi | Institute for Molecular Medicine Finland (FIMM), HiLIFE, University of Helsinki, Helsinki, Finland; Hospital District of Helsinki and Uusimaa, Helsinki, Finland | **Clinical Task Forces (FinnGen phase 3)** | **Pulmonology Task Force and fibrotic diseases interest group** |
| Raisa Serpi | Northern Finland Biobank Borealis / University of Oulu / Wellbeing services county of North Ostrobothnia, Oulu, Finland | **Clinical Task Forces (FinnGen phase 3)** | **Pulmonology Task Force and fibrotic diseases interest group** |
| Riitta Kaarteenaho | University of Oulu, Oulu, Finland | **Clinical Task Forces (FinnGen phase 3)** | **Pulmonology Task Force and fibrotic diseases interest group** |
| Hannu Kankaanranta | University of Gothenburg, Gothenburg, Sweden/ Seinäjoki Central Hospital, Seinäjoki, Finland/ Tampere University, Tampere, Finland | **Clinical Task Forces (FinnGen phase 3)** | **Pulmonology Task Force and fibrotic diseases interest group** |
| Coralie Viollet | Astra Zeneca, Cambridge, United Kingdom | **Clinical Task Forces (FinnGen phase 3)** | **Pulmonology Task Force and fibrotic diseases interest group** |
| Eleanor Wheeler | Astra Zeneca, Cambridge, United Kingdom | **Clinical Task Forces (FinnGen phase 3)** | **Pulmonology Task Force and fibrotic diseases interest group** |
| Oliver Burren | Astra Zeneca, Cambridge, United Kingdom | **Clinical Task Forces (FinnGen phase 3)** | **Pulmonology Task Force and fibrotic diseases interest group** |
| Christoph Ogris | Boehringer Ingelheim, Ingelheim am Rhein, Germany | **Clinical Task Forces (FinnGen phase 3)** | **Pulmonology Task Force and fibrotic diseases interest group** |
| Eric Simon | Boehringer Ingelheim, Ingelheim am Rhein, Germany | **Clinical Task Forces (FinnGen phase 3)** | **Pulmonology Task Force and fibrotic diseases interest group** |
| Frank LI | Boehringer Ingelheim, Ingelheim am Rhein, Germany | **Clinical Task Forces (FinnGen phase 3)** | **Pulmonology Task Force and fibrotic diseases interest group** |
| Julio Cesar Bolivar Lopez | Boehringer Ingelheim, Ingelheim am Rhein, Germany | **Clinical Task Forces (FinnGen phase 3)** | **Pulmonology Task Force and fibrotic diseases interest group** |
| Yao Hu | Boehringer Ingelheim, Ingelheim am Rhein, Germany | **Clinical Task Forces (FinnGen phase 3)** | **Pulmonology Task Force and fibrotic diseases interest group** |

| | | | |
|---|---|---|---|
| Zhihao Ding | Boehringer Ingelheim, Ingelheim am Rhein, Germany | **Clinical Task Forces (FinnGen phase 3)** | **Pulmonology Task Force and fibrotic diseases interest group** |
| Elena Sanchez | Bristol Myers Squibb, New York, NY, United States | **Clinical Task Forces (FinnGen phase 3)** | **Pulmonology Task Force and fibrotic diseases interest group** |
| Emily Holzinger | Bristol Myers Squibb, New York, NY, United States | **Clinical Task Forces (FinnGen phase 3)** | **Pulmonology Task Force and fibrotic diseases interest group** |
| Joe Maranville | Bristol Myers Squibb, New York, NY, United States | **Clinical Task Forces (FinnGen phase 3)** | **Pulmonology Task Force and fibrotic diseases interest group** |
| Lilith Moss | Bristol Myers Squibb, New York, NY, United States | **Clinical Task Forces (FinnGen phase 3)** | **Pulmonology Task Force and fibrotic diseases interest group** |
| Michael Turchin | Bristol Myers Squibb, New York, NY, United States | **Clinical Task Forces (FinnGen phase 3)** | **Pulmonology Task Force and fibrotic diseases interest group** |
| Zijie Zhao | Bristol Myers Squibb, New York, NY, United States | **Clinical Task Forces (FinnGen phase 3)** | **Pulmonology Task Force and fibrotic diseases interest group** |
| Diana Chang | Genentech, San Francisco, CA, United States | **Clinical Task Forces (FinnGen phase 3)** | **Pulmonology Task Force and fibrotic diseases interest group** |
| Audrey Chu | GlaxoSmithKline, Collegeville, PA, United States | **Clinical Task Forces (FinnGen phase 3)** | **Pulmonology Task Force and fibrotic diseases interest group** |
| Billy Fahy | GlaxoSmithKline, Collegeville, PA, United States | **Clinical Task Forces (FinnGen phase 3)** | **Pulmonology Task Force and fibrotic diseases interest group** |
| Jessica Chao | GlaxoSmithKline, Collegeville, PA, United States | **Clinical Task Forces (FinnGen phase 3)** | **Pulmonology Task Force and fibrotic diseases interest group** |
| Joanna Betts | GlaxoSmithKline, Collegeville, PA, United States | **Clinical Task Forces (FinnGen phase 3)** | **Pulmonology Task Force and fibrotic diseases interest group** |
| Jonathan Davitte | GlaxoSmithKline, Collegeville, PA, United States | **Clinical Task Forces (FinnGen phase 3)** | **Pulmonology Task Force and fibrotic diseases interest group** |
| Paola Bronson | GlaxoSmithKline, Collegeville, PA, United States | **Clinical Task Forces (FinnGen phase 3)** | **Pulmonology Task Force and fibrotic diseases interest group** |
| Prerak Desai | GlaxoSmithKline, Collegeville, PA, United States | **Clinical Task Forces (FinnGen phase 3)** | **Pulmonology Task Force and fibrotic diseases interest group** |
| Dermot Reilly | Johnson & Johnson Innovative Medicine, Spring House, PA, United States | **Clinical Task Forces (FinnGen phase 3)** | **Pulmonology Task Force and fibrotic diseases interest group** |
| Mona Selej | Johnson & Johnson Innovative Medicine, Spring House, PA, United States | **Clinical Task Forces (FinnGen phase 3)** | **Pulmonology Task Force and fibrotic diseases interest group** |
| P Dunnmon | Johnson & Johnson Innovative Medicine, Spring House, PA, United States | **Clinical Task Forces (FinnGen phase 3)** | **Pulmonology Task Force and fibrotic diseases interest group** |
| Jorge Del-aguila | Merck, Kenilworth, NJ, United States | **Clinical Task Forces (FinnGen phase 3)** | **Pulmonology Task Force and fibrotic diseases interest group** |
| Jozsef Karman | Merck, Kenilworth, NJ, United States | **Clinical Task Forces (FinnGen phase 3)** | **Pulmonology Task Force and fibrotic diseases interest group** |
| Travis Barr | Merck, Kenilworth, NJ, United States | **Clinical Task Forces (FinnGen phase 3)** | **Pulmonology Task Force and fibrotic diseases interest group** |
| Katherine Mccauley | Novartis Institutes for BioMedical Research, Cambridge, MA, United States | **Clinical Task Forces (FinnGen phase 3)** | **Pulmonology Task Force and fibrotic diseases interest group** |
| Xiaobo Xia | Novartis Institutes for BioMedical Research, Cambridge, MA, United States | **Clinical Task Forces (FinnGen phase 3)** | **Pulmonology Task Force and fibrotic diseases interest group** |
| Madhurima Saxena | Pfizer, New York, NY, United States | **Clinical Task Forces (FinnGen phase 3)** | **Pulmonology Task Force and fibrotic diseases interest group** |

| | | | |
|---|---|---|---|
| Pitchumani Sivakumar | Pfizer, New York, NY, United States | **Clinical Task Forces (FinnGen phase 3)** | **Pulmonology Task Force and fibrotic diseases interest group** |
| Sumedha Jassal | Pfizer, New York, NY, United States | **Clinical Task Forces (FinnGen phase 3)** | **Pulmonology Task Force and fibrotic diseases interest group** |
| David Habiel | Translational Sciences, Sanofi R&D, Framingham, MA, USA | **Clinical Task Forces (FinnGen phase 3)** | **Pulmonology Task Force and fibrotic diseases interest group** |
| Guanling Huan | Translational Sciences, Sanofi R&D, Framingham, MA, USA | **Clinical Task Forces (FinnGen phase 3)** | **Pulmonology Task Force and fibrotic diseases interest group** |
| Marika Kaakinen | Institute for Molecular Medicine Finland (FIMM), HiLIFE, University of Helsinki, Helsinki, Finland | **Clinical Task Forces (FinnGen phase 3)** | **Parkinson´s disease Task Force** |
| Mary Pat Reeve | Institute for Molecular Medicine Finland (FIMM), HiLIFE, University of Helsinki, Helsinki, Finland; Broad Institute, Cambridge, MA, United States | **Clinical Task Forces (FinnGen phase 3)** | **Parkinson´s disease Task Force** |
| Filip Scheperjans | Hospital District of Helsinki and Uusimaa, Helsinki, Finland | **Clinical Task Forces (FinnGen phase 3)** | **Parkinson´s disease Task Force** |
| Andrew Blumenfeld | AbbVie, Chicago, IL, United States | **Clinical Task Forces (FinnGen phase 3)** | **Parkinson´s disease Task Force** |
| Britney Milkovich | AbbVie, Chicago, IL, United States | **Clinical Task Forces (FinnGen phase 3)** | **Parkinson´s disease Task Force** |
| Jan Freudenberg | AbbVie, Chicago, IL, United States | **Clinical Task Forces (FinnGen phase 3)** | **Parkinson´s disease Task Force** |
| Tushar Kumar | AbbVie, Chicago, IL, United States | **Clinical Task Forces (FinnGen phase 3)** | **Parkinson´s disease Task Force** |
| Hans van Leeuwen | Bayer AG, Leverkusen, Germany | **Clinical Task Forces (FinnGen phase 3)** | **Parkinson´s disease Task Force** |
| Juho Immonen | Bayer AG, Leverkusen, Germany | **Clinical Task Forces (FinnGen phase 3)** | **Parkinson´s disease Task Force** |
| Samu Kurki | Bayer AG, Leverkusen, Germany | **Clinical Task Forces (FinnGen phase 3)** | **Parkinson´s disease Task Force** |
| Coro Paisan-Ruiz | Biogen, Cambridge, MA, United States | **Clinical Task Forces (FinnGen phase 3)** | **Parkinson´s disease Task Force** |
| Anna Podgornaia | Bristol Myers Squibb, New York, NY, United States | **Clinical Task Forces (FinnGen phase 3)** | **Parkinson´s disease Task Force** |
| Benjamin Sun | Bristol Myers Squibb, New York, NY, United States | **Clinical Task Forces (FinnGen phase 3)** | **Parkinson´s disease Task Force** |
| Janie Shelton | Bristol Myers Squibb, New York, NY, United States | **Clinical Task Forces (FinnGen phase 3)** | **Parkinson´s disease Task Force** |
| Peng Jiang | Bristol Myers Squibb, New York, NY, United States | **Clinical Task Forces (FinnGen phase 3)** | **Parkinson´s disease Task Force** |
| Stephanie Loomis | Bristol Myers Squibb, New York, NY, United States | **Clinical Task Forces (FinnGen phase 3)** | **Parkinson´s disease Task Force** |
| Tushar Bhangale | Genentech, San Francisco, CA, United States | **Clinical Task Forces (FinnGen phase 3)** | **Parkinson´s disease Task Force** |
| John Eicher | GlaxoSmithKline, Collegeville, PA, United States | **Clinical Task Forces (FinnGen phase 3)** | **Parkinson´s disease Task Force** |
| Abolfazl Doostparast Torshizi | Johnson & Johnson Innovative Medicine, Spring House, PA, United States | **Clinical Task Forces (FinnGen phase 3)** | **Parkinson´s disease Task Force** |
| Aristide Merola | Merck, Kenilworth, NJ, United States | **Clinical Task Forces (FinnGen phase 3)** | **Parkinson´s disease Task Force** |

| | | | |
|---|---|---|---|
| Oliver Freeman | Merck, Kenilworth, NJ, United States | **Clinical Task Forces (FinnGen phase 3)** | **Parkinson´s disease Task Force** |
| Simonne Longerich | Merck, Kenilworth, NJ, United States | **Clinical Task Forces (FinnGen phase 3)** | **Parkinson´s disease Task Force** |
| Mari Niemi | Novartis Institutes for BioMedical Research, Cambridge, MA, United States | **Clinical Task Forces (FinnGen phase 3)** | **Parkinson´s disease Task Force** |
| Katherine Klinger | Translational Sciences, Sanofi R&D, Framingham, MA, USA | **Clinical Task Forces (FinnGen phase 3)** | **Parkinson´s disease Task Force** |
